# Intestinal somatic mutations in inflammatory bowel disease patients are enriched in very early onset IBD and primary immunodeficiency genes

**DOI:** 10.1101/2024.12.09.24318484

**Authors:** Iwan J. Hidding, Arnau Vich Vila, Shixian Hu, Arno R. Bourgonje, Marielle van Gijn, W.T.K. Maassen, Gerben B. van der Vries, Renate A.A.A. Ruigrok, Sergio Andreu-Sánchez, Bernadien H. Jansen, Hendrik M. van Dullemen, Marijn C. Visschedijk, Gerard Dijkstra, Morris A. Swertz, Cleo C. van Diemen, Eleonora A.M. Festen, Rinse K. Weersma, Floris Imhann

## Abstract

The genetic predisposition of Inflammatory bowel disease (IBD) in the germline is well-established, while the potential role of somatic mutations in IBD remains underexplored. We aim to identify somatic mutations in intestinal mucosa and study their potential role in IBD pathogenesis. We identified somatic mutations using 538 intestinal biopsies and 268 blood samples from 268 IBD patients and 51 intestinal biopsies from 19 non-IBD controls. Mutations were called from RNA-seq data and whole exome sequencing data using Mutect2 software. Pathogenicity of mutations was predicted using the Variant Interpretation Pipeline. Mutation frequency and patterns were compared between IBD and controls, and within IBD-subgroups, considering both inflammation status and tissue of origin. IBD-affected tissue exhibited a significantly higher burden of somatic mutations compared to controls (p=0.02), with a striking enrichment for predicted likely pathogenic or pathogenic (pLP-P) mutations (p=2.07e-09). Notably, within IBD biopsies, pLP-P mutations were enriched in very early onset-IBD (VEO-IBD) genes (p≤2.2e-16) and primary immune deficiency (PID) genes (p≤2.2e-16), showing a 5.5-fold increase compared to non-IBD. Inflamed tissue showed an increase in mutations in PID and IBD-GWAS associated genes. Additionally, we identified pathogenic mutations in many IBD genes, such as the stop-gained mutation (c.517C>T) in *HPS1*, and many pLP-P mutations across the JAK-STAT pathway. Our analysis reveals widespread accumulation of pLP-P somatic mutations in IBD-affected tissue compared to non-IBD controls, with significant enrichment in genes that cause monogenic VEO-IBD. These findings suggest that somatic mutations can contribute to the initiation and perpetuation of inflammation in IBD.

## Introduction

Inflammatory bowel disease (IBD) is a relapsing-remitting disorder characterized by mucosal inflammation in the gastrointestinal tract. IBD is a common disorder, affecting 0.5–1% of the European population (2.5–3 million individuals)[1]. The exact etiology and pathogenesis of IBD are not yet fully understood, but it is considered to be an exaggerated immune response to gut microbiota that has been altered in a pro-inflammatory manner by environmental triggers, occurring in a genetically predisposed host[2–5]. Genome-wide-association studies (GWAS) and whole exome sequencing (WES) studies have partially unraveled the genetic predisposition to IBD in the germline[6,7]. GWAS have so far detected 320 common risk loci and several rare variants in IBD-related genes such as *NOD2*[6]. However, known genetic factors currently only explain about 6.5% of IBD risk[6].

A role for somatic mutations in the pathogenesis of IBD was recently proposed[8,9]. Somatic mutations are DNA mutations not present in the germline at conception. They are acquired later in life through random replication errors and environmental stressors such as exposure to chemical compounds and free radicals. Once introduced, somatic mutations can spread through normal cell division[8]. Because of this mechanism, these mutations are only present locally in the affected tissue and often cannot be detected when sequencing DNA from blood.

Somatic mutations are known to be one of the causes of cancer. For example, somatic mutations in the *TP53*, *APC*, *KRAS* and *BRAF* genes in colonic epithelial cells can induce cell proliferation and promote cell survival, leading to colorectal cancer[10–13]. The role of somatic mutations in the pathogenesis of non-cancer diseases is less well-studied. In IBD, somatic mutations in *NFKBIZ*, *ZC3H12A* and *PIGR* have been observed to accumulate in gut epithelial tissue of ulcerative colitis (UC) patients. These mutations have been shown to confer resistance to the IL-17A pro-apoptotic response[9] that is common in IBD, demonstrating their potential impact on disease. However, the role of somatic mutations in IBD could be much larger since there are several mechanisms that can increase the occurrence and impact of somatic mutations in the gut. First, gut tissue is prone to accumulation of somatic mutations in stem cells due to the rapid, constant clonal cell division that replenishes the epithelium of the villi, leading to accumulation of 40 mutations per crypt per year[8]. Second, during chronic inflammation, many free radicals are produced within cells, leading to additional DNA damage and somatic mutations, estimated at 55 additional mutations per crypt per year[8]. Third, gut inflammation can lead to an increased clonal crypt size[8], enhancing the impact of somatic mutations. Finally, studies have shown that certain microbiota can induce specific somatic mutations in the gut[14,15].

We hypothesized that we could call somatic mutations using biopsy-derived RNA-sequencing (RNAseq) and determine if these mutations were enriched in specific IBD-related gene sets and related to IBD sub-phenotypes. To test these hypotheses, we analyzed somatic mutations by investigating genotype-calling inconsistencies between RNAseq data from 538 intestinal biopsies and WES data from blood in a cohort of 268 IBD patients and compared them with RNAseq data from 51 intestinal biopsies from 19 non-IBD controls. We focused particularly on somatic mutations in genes in IBD-associated GWAS loci and those in genes in which defects can cause monogenic very early onset IBD (VEO-IBD) and monogenic primary immune deficiencies (PIDs).

## Methods

### Cohort description and institutional review board approval

This study included data from 268 IBD patients and 19 non-IBD control participants from the University Medical Center Groningen (UMCG) who are included in the 1000IBD biobank (1000IBD UMCG IRB no. 2008.338)[16]. Prior to sample collection, all participants provided written informed consent. See **Online supplementary methods** for a detailed cohort description. A full schematic overview of the methods is given in **Figure 1**. All UMCG IDs in the **Supplemental tables** are anonymized research ids only known to the research group.

**Figure 1.**
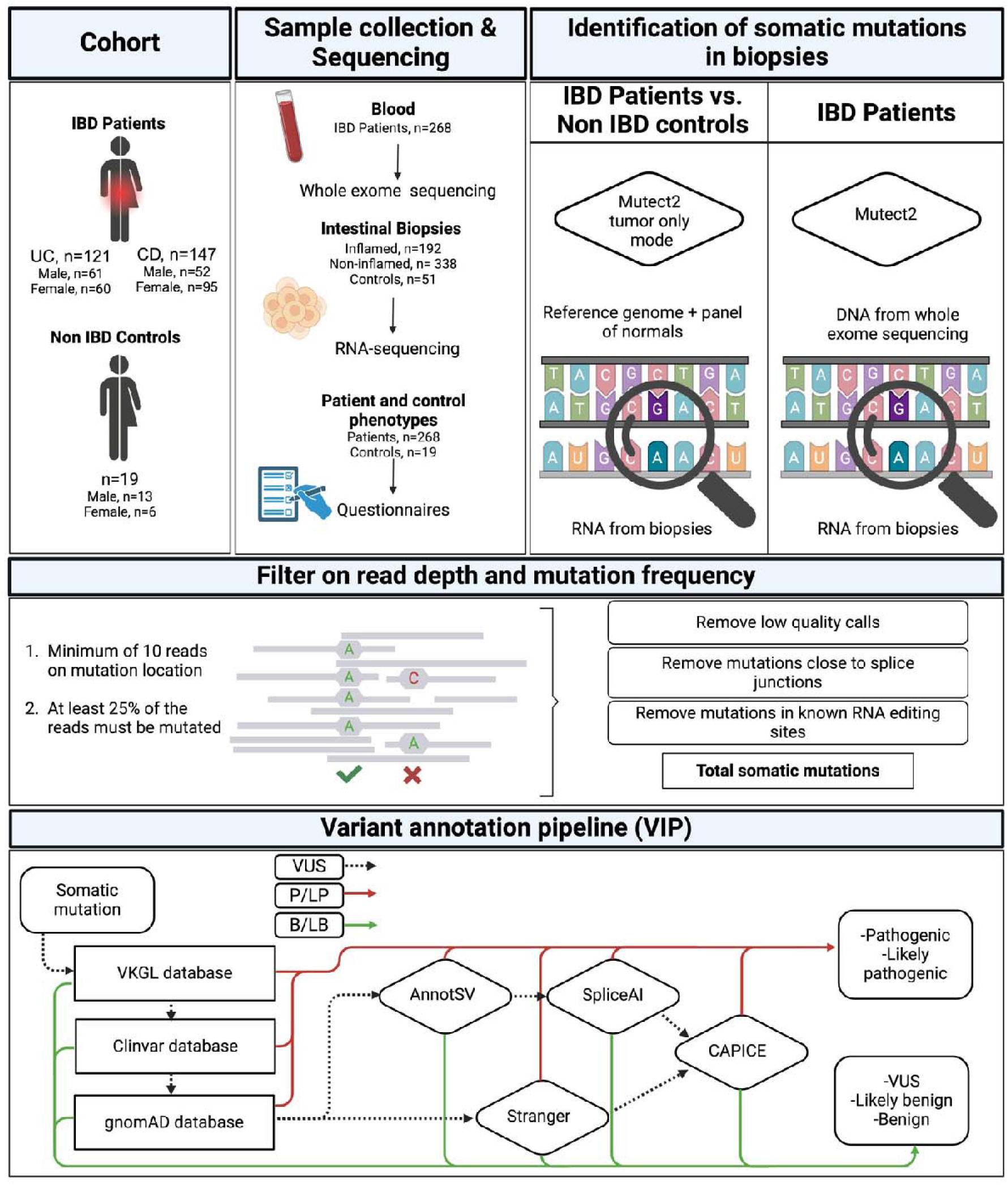
Study cohort and methods. Samples and data were collected from 268 Inflammatory Bowel Disease patients (Crohn’s disease (CD) n=147, ulcerative colitis (UC) n=121) and 19 non-IBD control participants. Multiple intestinal biopsies were taken from inflamed (IBD n=192) and non-inflamed (IBD n=338, controls n=51) regions. Blood was collected from each IBD patient, and IBD patient phenotypes were recorded at colonoscopy and at outpatient clinic appointments. RNA-sequencing (RNAseq) was performed on all biopsies. To compare somatic mutations between IBD patients and controls, Mutect2 tumor-only mode was used to call somatic mutations in the RNAseq data. For IBD patients, whole exome sequencing (WES) data was available, so both WES and RNAseq data were used to call somatic mutations. Extensive filtering was applied based on minimum read depth and variant allele frequency. Low quality mutation calls, mutations close to splice junctions and mutations in RNA-editing sites were removed as they might not reliably represent somatic mutations in the DNA. The somatic mutations identified were classified, using the Variant Interpretation Pipeline (VIP) with the default configuration, according to American College of Medical Genetics guidelines: Class 1 Benign (B), Class 2 Likely Benign (LB), Class 3 Variant of Unknown Significance (VUS), Class 4 Likely Pathogenic (LP), Class 5 Pathogenic (P).

### Clinical data

During every outpatient department visit, clinical data of IBD patients is collected by the treating gastroenterologist and stored in the electronic health record, then subsequently exported to the 1000IBD database[17]. For this study, we extracted pseudonymized clinical data from the 1000IBD database (see **Table 1** for an overview of clinical characteristics).

**Table 1.**
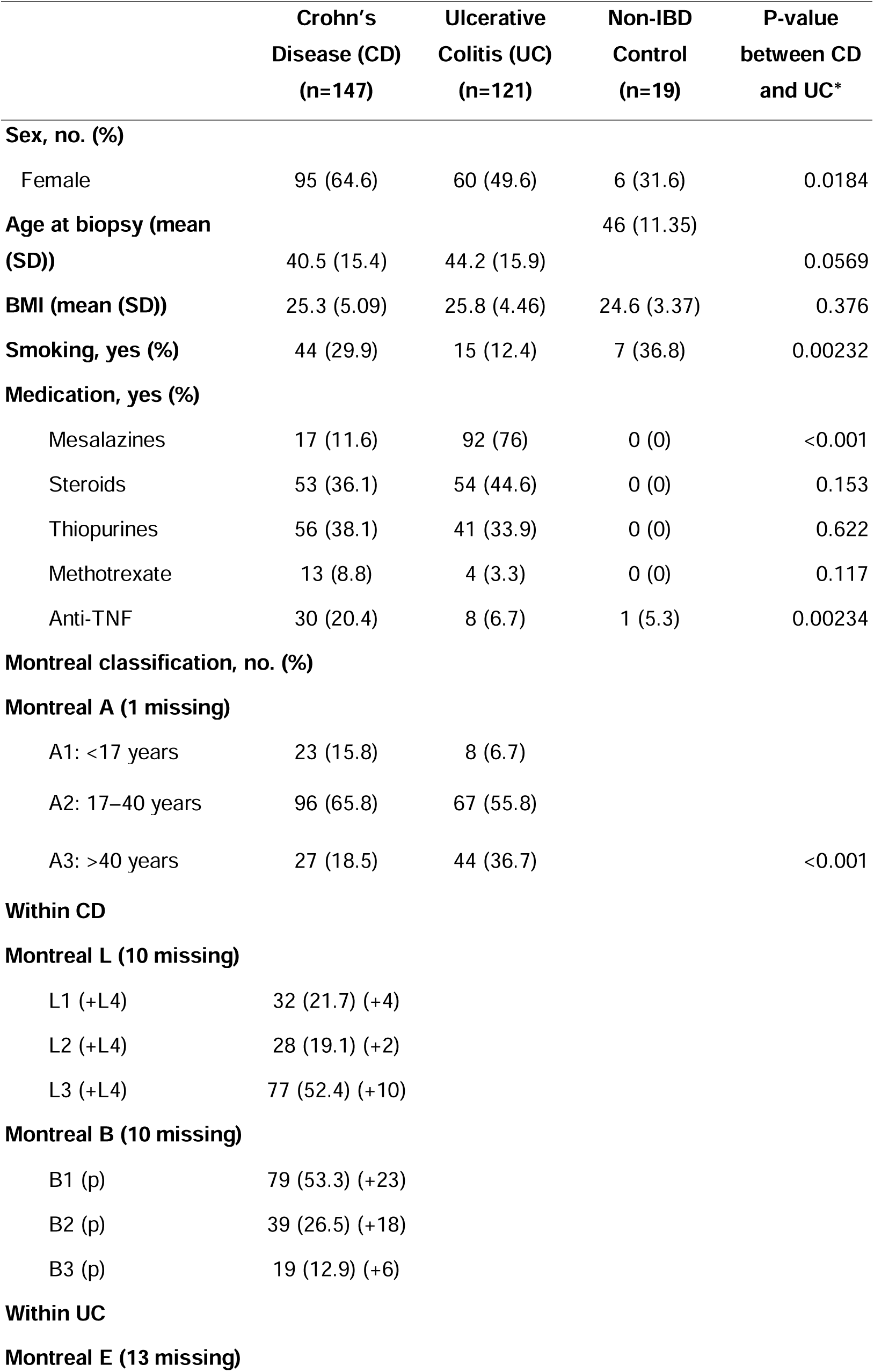
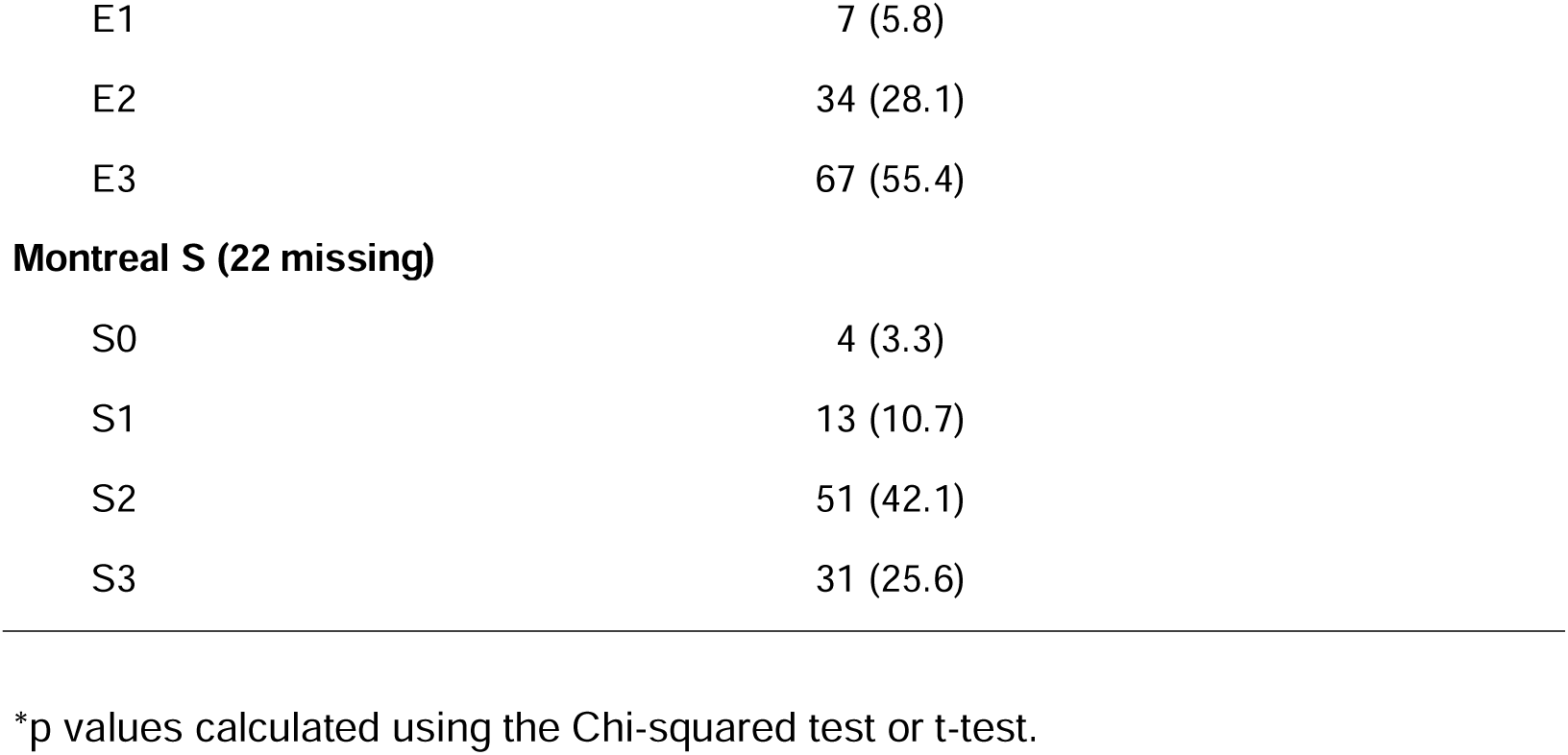
Cohort description.

### Biopsy and blood sample collection and storage

From 268 IBD participants, 589 intestinal mucosal biopsies were collected during colonoscopy procedures (see **Supplementary table S1** for an overview). For each IBD participant, at least one inflamed biopsy and 0–5 non-inflamed biopsies were collected. From the 19 non-IBD participants, 51 biopsies were collected from non-inflamed locations. Biopsies were immediately snap-frozen on-site and stored at -80℃ until processing. Biopsy location and macroscopic inflammation status were registered by the gastroenterologist during colonoscopy. Biopsies were also assessed by an independent pathologist to confirm the inflammation status.

### RNA isolation from intestinal biopsies and RNAseq

RNA was isolated from gut biopsies using the AllPrep DNA/RNA mini kit from Qiagen according to the manufacturer’s instructions, as previously described[18]. Paired-end RNAseq was performed in two batches on the Illumina NextSeq500 and the Illumina HiSeq PE250 platforms. See the **Online supplementary methods** for full details.

### DNA isolation from blood and whole exome sequencing

Blood was collected from 268 IBD participants. DNA isolation was performed using the AutoPure LS procedure from Qiagen. WES was performed at the Broad Institute of Harvard and MIT (Boston, USA) using the Illumina HiSeq 2500 platform, as previously described[18,19], generating 86.06 million high quality reads per sample on average. WES data is not available for the non-IBD controls. Full details are provided in the **Online supplementary methods**.

### Prediction and identification of somatic mutations

To compare the number of somatic mutations between biopsies taken from IBD patients and biopsies taken from non-IBD controls, we used GATK (v4.2.4.1) Mutect2 in tumor-only mode, a reference panel from GATK (1000g_pon.hg38.vcf.gz) containing potential recurrent technical artifacts, and the hg38 reference genome. Tumor-only mode enables the prediction of somatic mutations based on RNAseq data only (i.e. without blood WES data). We followed GATK best practices for variant discovery[20], aligning the RNAseq using STAR aligner (v2.7.3a)[21] and then using MarkDuplicatesSpark and BaseRecalibrator to reduce technical artifacts. Post-alignment files were the input for Mutect2. To compare somatic mutations in the IBD patient biopsies between sub-phenotypes, we followed the same steps using matched WES data of the IBD patients rather than the reference panel. See **Online supplementary methods** for more details.

### Quality control: filtering

To reduce artifacts and sequencing errors, we performed stringent filtering after calling the somatic mutations (see **Online supplementary methods** for filtering steps and the number of mutations that were filtered out). Using FilterMutectCalls with standard settings, we removed low quality and low confidence calls based on the probability of technical artifacts, non-somatic and sequencing errors. Moreover, only somatic mutations with >25% variant allele frequency (VAF) on parts of the exome covered by at least 10 reads were used in further analyses. Finally, we removed mutations close to splice sites and mutations in known RNA-editing sites (both known to be unreliable in RNAseq data) using the RADAR and DARNED databases[22–24]. After filtering, we retained an average of 743 high confidence somatic mutations per biopsy for further analyses.

### Mutation annotation using the MOLGENIS Variant Interpretation Pipeline

To interpret somatic mutations, we used the Variant Interpretation Pipeline (VIP) created by MOLGENIS,[17,25]. This is a pipeline of “best-in-practice” tools for variant annotation and classification prediction, including the machine-learning-based variant pathogenicity predictor CAPICE (v5.1.2)[26]. VIP supports classification of mutations according to American College of Medical Genetics (ACMG) guidelines into the following classes: Class 1 Benign (B), Class 2 Likely Benign (LB), Class 3 Variant of Unknown Significance (VUS), Class 4 Likely Pathogenic (LP) and Class 5 Pathogenic (P)[27]. By default, VIP checks the Clinvar and Dutch national variant VKGL databases for known mutations. Subsequently, we also used VIP to check gnomAD for population frequency and evolutionary preservation[28]. For structural and splice variations, we used annotSV and then SpliceAI, with each tool able to call any variant as LP or not. When none of the above tools indicated a LP variant, a CAPICE score between 0 (predicted to be B) and 1 (predicted to be P) was used for the final classification. When a mutation was not previously registered as P in Clinvar or VKGL, the prediction algorithm classified the pathogenicity only as high as LP, following ACMG classification guidelines[27].

### Prioritizing genes using four IBD-related gene sets

To prioritize genes of interest, we used four gene sets: 103 genes with mutations known to cause monogenic VEO-IBD that is used as a diagnostic panel in the UMCG Clinical Genetics Laboratory, 475 monogenic PID genes also used as a diagnostic panel, 318 GWAS Candidate genes identified in the 320 IBD-associated loci by Liu et al[6] and 3504 GWAS Loci genes considering all genes in the loci windows based on LD blocks identified by Liu et al (see **Supplementary tables S2, S3, S4** and **S5** for full gene lists). Overlap between the gene sets is shown in **supplemental figure S1**. In total, 79 genes overlap between the PID and VEO-IBD gene sets, 13 overlap between the GWAS Candidate genes and VEO-IBD gene sets, and 44 overlap between the PID and GWAS Candidate gene sets. All LP and P variants in these gene sets were reviewed by expert clinical laboratory geneticist MvG to confirm the classification.

### Comparing mutations in IBD patient biopsies to non-IBD controls

To compare the number of somatic mutations found between IBD patients and non-IBD controls, we counted the total number of somatic mutations per biopsy and analyzed the effect of the disease using linear models correcting for inflammation, biopsy location, sex, age at biopsy and participant ID. We then counted the number of somatic mutations in each of the four gene sets for each participant. Here again, we analyzed the effect of the disease using linear models correcting for inflammation, biopsy location, sex, age at biopsy and participant ID. These analyses were performed on both the total number of somatic mutations (all classes from B to P) and on mutations that potentially cause disease (LP and P only).

### Somatic mutation enrichment in IBD-related gene panels in biopsies of IBD patients

We tested whether there are more somatic mutations in the four IBD-related gene sets compared to the rest of the exome (called from transcriptome) in biopsies of IBD patients. To do so, we summed all the somatic mutations in each gene set for every biopsy of an IBD patient and divided this number by the gene length of all the genes in the gene set, correcting for both the number of genes per gene set and the length of the genes based on Refseq reference length (see formula), as the odds of any random mutation occurring in a gene increases with gene length[29].

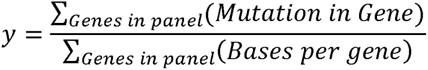

Differences between the number of somatic mutations in a gene set and the 16,688 non-IBD-related Refseq genes (all 20,078 protein-coding Refseq genes minus the genes in the four panels) were analyzed using Wilcoxon signed-rank tests. These analyses were performed on both the total number of somatic mutations (all classes from B to P) and the mutations that potentially cause disease (LP and P).

### Statistical analyses

As a somatic signatures in colorectal cancer genes can be expected when calling somatic mutations in the intestine, we analyzed the relationship between age and the number of somatic mutations per biopsy in 12 known colorectal cancer genes (*APC, TP53, TTN, KRAS, MUC16, SYNE1, FAT4, PIK3CA, OBSCN, ZFHX4, RYR2* and *CSMD3*). This analysis was performed using linear models for all somatic mutations (all classes from B to P) and for mutations that potentially cause disease (LP and P). We also tested the influence of the inflammation status of the tissue at the time of biopsy, correcting for biopsy location and sequencing batch. See **Online supplementary methods** for more details.

### Data online

Data are uploaded to the European Genome-Phenome Archive (EGA) and are available upon request. The data for the Groningen IBD cohort can be requested with EGA the accession number EGAS00001002702 (https://ega-archive.org/).

### Code online

All code in Python and R is available at https://github.com/GRONINGEN-MICROBIOME-CENTRE/Somatic_mutations_IBD.

## Results

### Gut biopsies of IBD patients show significantly higher accumulation of somatic mutations

We called 621,471 somatic mutations in the RNA of 436 gut biopsies from 256 IBD patients and 55,480 somatic mutations in the RNA of 51 gut biopsies from 19 non-IBD controls, using only the IBD biopsies that were sequenced in the same batch as the control biopsies. We first tested the association between the number of somatic mutations detected and various clinical phenotypes. Here, we observed no significant difference in the number of mutations when considering sex, age, IBD subtype, smoking status, or location of the biopsy. We did observe a negative association with disease duration (Pearson correlation, p=5.27e-05) and a significant increase in somatic mutations in inflamed biopsies (p=0.005). After correcting for these clinical characteristics, we observed a statistically significant increase in the total number of somatic mutations in biopsies from IBD patients compared to biopsies from non-IBD controls (p=0.02) (**Figure 2A**). After annotating the somatic mutations for pathogenicity using VIP, we looked only at potentially disease-causing mutations (LP and P) and observed a much larger difference between IBD patients and non-IBD controls (p=2.07e-09) (**Figure 2B**).

**Figure 2.**
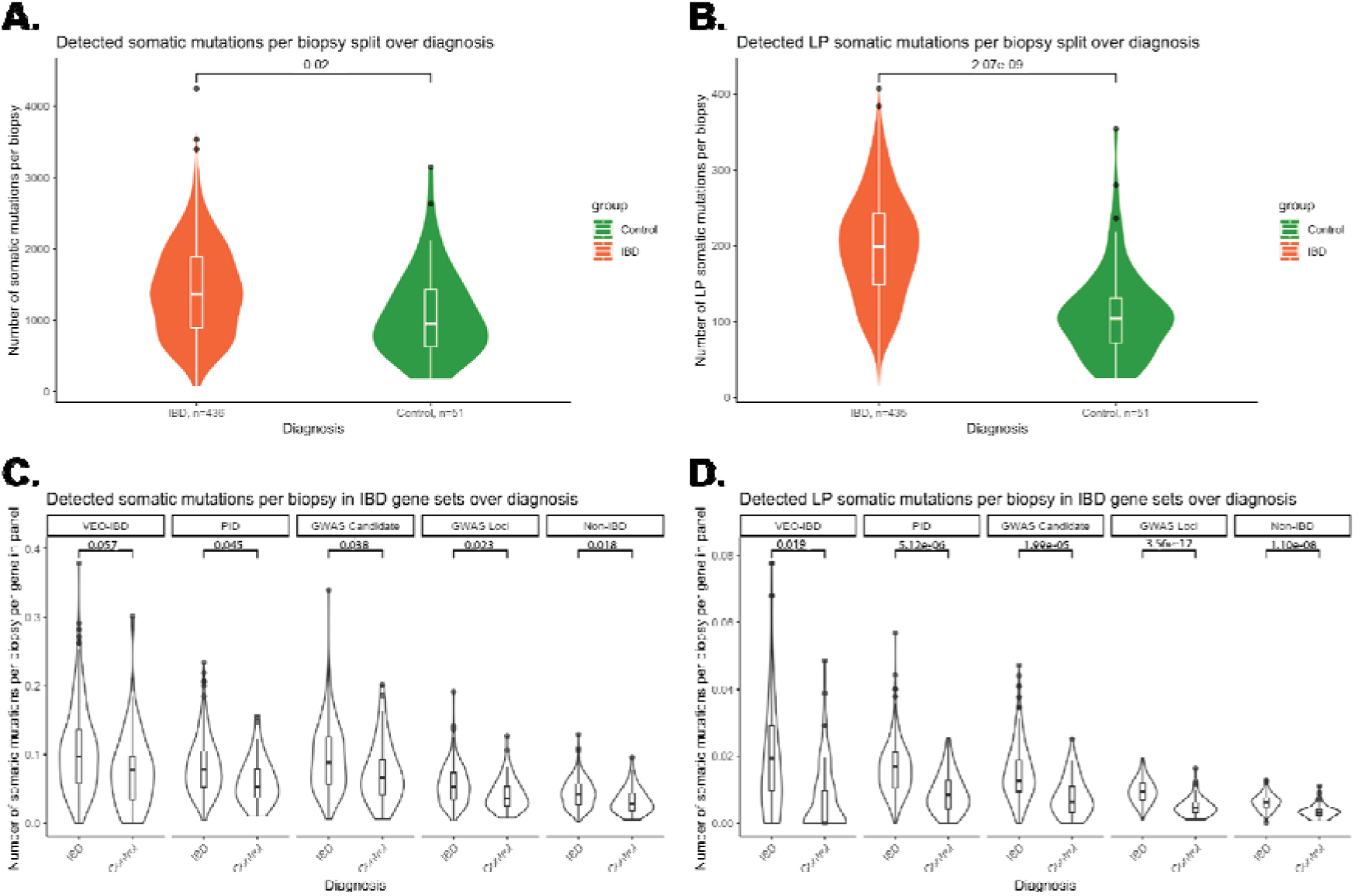
Increased number of somatic mutations in IBD intestinal biopsies compared to non-IBD controls s. **A)** Number of somatic mutations detected in biopsies of IBD patients (orange) and non-IBD controls (green). Mutations were called using Mutect2 in tumor-only mode. A significant difference is observed between IBD patients and non-IBD controls (p=0.02). **B)** Number of potentially disease-causing LP and P mutations detected. The difference between IBD patients (orange) and non-IBD controls (green) is larger when comparing only LP–P somatic mutations instead of all somatic mutations (p=2.07e-09). **C)** Number of somatic mutations detected per gene in four gene panels of IBD-associated genes (see **Supplementary tables 2– 5**). A statistically significantly increased accumulation of somatic mutations (Classes 1–5) is observed in IBD patients compared to non-IBD controls for all gene panels except the Very Early Onset-IBD (VEO-IBD) genes (VEO-IBD p=0.057, Primary Immune Deficiency (PID) p=0.045, GWAS Candidate p=0.038, GWAS Loci p=0.023, non-IBD p=0.018). **D)** Number of potentially disease-causing LP-P somatic mutations detected in the four gene panels. For all gene panels, an increased accumulation of LP–P somatic mutations is observed in IBD patients compared to non-IBD controls (VEO-IBD p=0.019, PID p=5.12e-06, GWAS Candidate p=1.99e-05, GWAS Loci p=3.56e-12, non-IBD p=1.10e-08). Again, the difference between IBD patients and non-IBD controls is larger when looking at potentially disease-causing somatic mutations. P values were calculated using linear models correcting for age, sex, BMI, location and inflammation status of the biopsy.

### IBD-associated genes accrue more somatic mutations in IBD patients compared to controls

We prioritized four gene sets containing genes relevant to IBD pathogenesis. These gene sets contain VEO-IBD genes (n=103), PID genes (n=475), GWAS Candidate genes (n=318) and GWAS Loci genes (n=3504) (see **Supplementary tables S2–S5**). A comparison of the total number of somatic mutations (Classes B–P) in each gene set between IBD patients and non-IBD controls revealed that the accumulation of somatic mutations was significantly larger in all but the VEO-IBD gene sets in IBD patients after correcting for age, sex, BMI, inflammation status and location of biopsy origin ( VEO-IBD p=0.057, PID p=0.045, GWAS Candidate p=0.038, GWAS Loci p=0.023, non-IBD-associated genes p=0.018). Comparing only potentially disease-causing somatic mutations (P and LP) showed an even larger accumulation in IBD patients, including a near-absence of potentially disease-causing mutations in VEO-IBD genes in the non-IBD control biopsies (VEO-IBD p=0.019, PID p=5.12e-06, GWAS Candidate p=1.99e-05, GWAS Loci p=3.56e-12, non-IBD p=1.10e-08) (**Figures 2C, D**). We observed no increase in somatic mutations with age in all genes or in only the 12 colorectal cancer genes, considering only one sample per patient taken at random (Pearson correlation, Age: corr=-0.110, p=0.063, Colorectal genes: corr=-0.003, p=0.62; **Supplementary figures S2, S3**). We also observed a significant increase in somatic mutations in inflamed biopsies compared to non-inflamed biopsies after VIP annotation and pLP-P selection (Mann-Whitney U, all mutations p=0.001, P/LP mutations p=3.72e-06; **Supplementary figures S4, S5**).

### IBD patients accumulate more somatic mutations in VEO-IBD genes than non-IBD controls

We then used the WES data (where available) to filter out somatic mutations. Applying the same filtering steps as before, we called 400,544 somatic mutations in 538 gut biopsies from 268 IBD patients (91,536 LP-P mutations) (**Supplementary figure S6**). We analyzed the enrichment of somatic mutations in the previously described gene sets within IBD biopsies. Here we observed a significant increase in VEO-IBD and PID genes compared to non-IBD genes (Wilcoxon signed-rank, VEO-IBD: 4.5-fold, p≤2.2e-16; PID: 1.6-fold, p≤2.2e-16). A significant decrease was observed in the GWAS Candidate and GWAS Loci genes (Wilcoxon signed-rank, Candidate: 0.8-fold, p≤2.2e-16; Loci: 0.9-fold, p≤2.2e-16). After pLP-P annotation, these effects remained the same (Wilcoxon signed-rank, VEO-IBD: 5.5-fold, p≤2.2e-16; PID: 1.5-fold, p≤2.2e-16; Candidate: 0.7-fold, p≤2.2e-16; Loci: 0.9-fold, p=3.80e-10) (**Figure 3A, B**). Additionally, we observed a significant enrichment in VEO-IBD and PID genes of mutations found in the Clinvar database and an enrichment in the autosomal dominant and X-linked mutations found in male patients in the PID gene panel (chi-squared, Clinvar: p=2.88e-87, inheritance: p=3.15e-139) (**Supplementary tables S6, S7**). Comparing the inflammation status over the different gene panels, we observed a significant enrichment under inflamed conditions in PID, GWAS Loci and GWAS Candidate genes, but not in VEO-IBD genes (**Figure 3C**).

**Figure 3.**
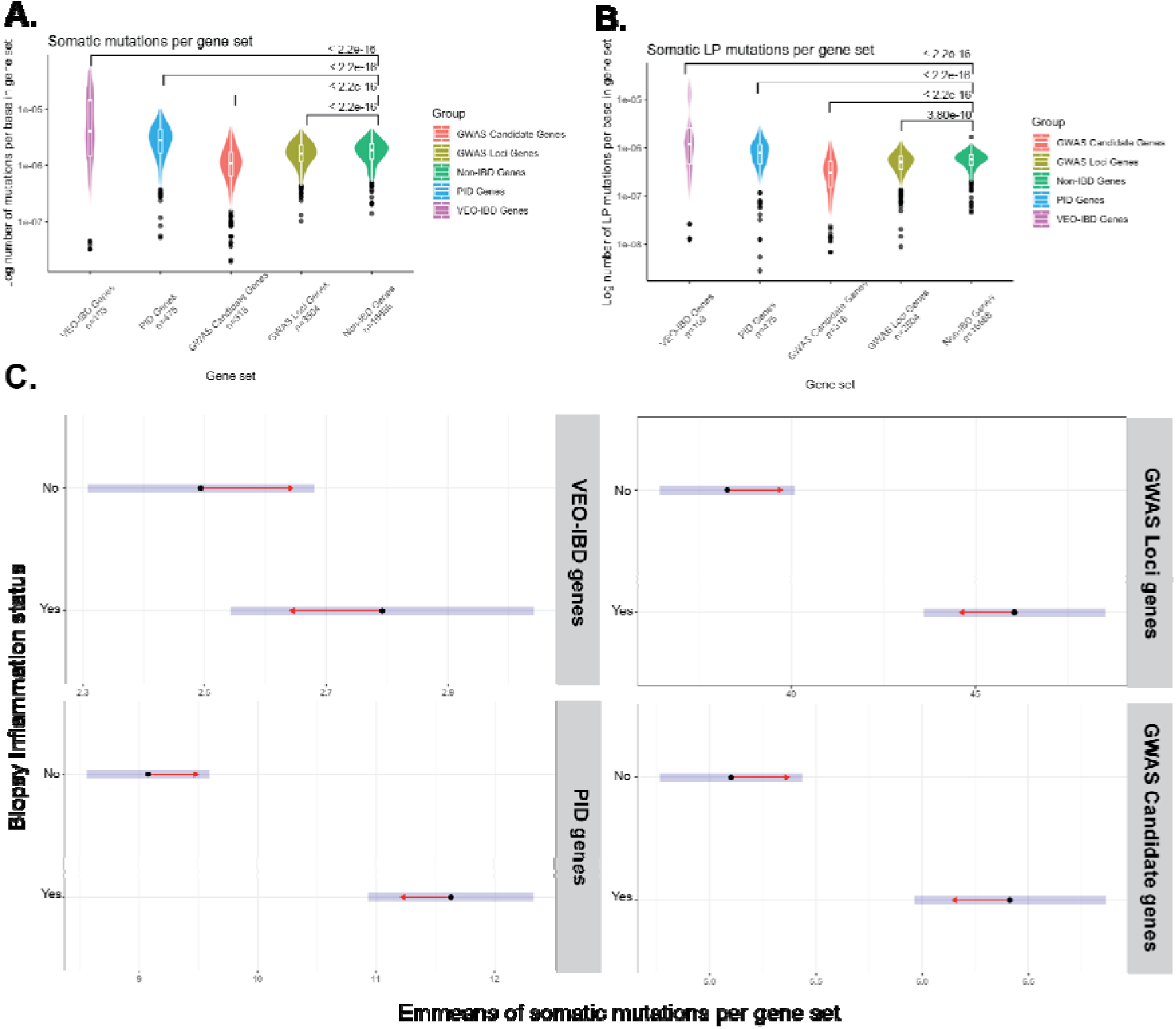
Gene set and inflammation status enrichment within IBD biopsies. **A**) Number of somatic mutations per gene set, corrected for gene length and number of genes within each gene set. We observe a significant accumulation of somatic mutations for Very Early Onset-IBD (VEO-IBD) and Primary immune deficiency (PID) genes compared to non-IBD genes (Wilcoxon signed-rank, VEO-IBD p≤2.2e-16, PID p≤2.2e-16). **B**) Number of somatic mutations per gene set after P/LP annotation using the VIP, corrected for the gene length and number of genes within each gene set. A significant accumulation of somatic mutations is observed for VEO-IBD and PID genes compared to non-IBD genes (Wilcoxon signed-rank, VEO-IBD p≤2.2e-16, PID p≤2.2e-16). **C**) Overview of somatic mutation accumulation based on biopsy inflammation status at time of sampling. Data is corrected for biopsy location, sequencing batch and patient ID (in case of multiple samples per patient) using linear models. Shown are the estimated marginal means (Emmeans) comparing the number of somatic mutations found per gene set over biopsy inflammation status at time of sampling. Arrows indicate the direction of the comparison. Significance is indicated by non-overlapping estimated means. A significant difference is observed in GWAS Loci genes, PID genes and GWAS Candidate genes, but not in VEO-IBD genes.

### Accumulation of LP mutations in functional domains in genes involved in IBD pathogenesis

As we identified 91,536 pLP-P somatic mutations in IBD biopsies, we sorted these mutations based on their associated inheritance and gene set and limited our subsequent analysis to a few key IBD genes. See **Supplemental tables S8-S35** for a full overview of the VIP- and Clinvar-annotated somatic mutations, sorted by gene set, database annotation and inheritance type.

Then, to understand the possible effect of pLP-P somatic mutations, we selected eight genes for in-depth analysis. These genes were chosen based on their relevance to IBD and the identified mutations (see **Figure 4** for genes and their functional domains and mutations).

**Figure 4.**
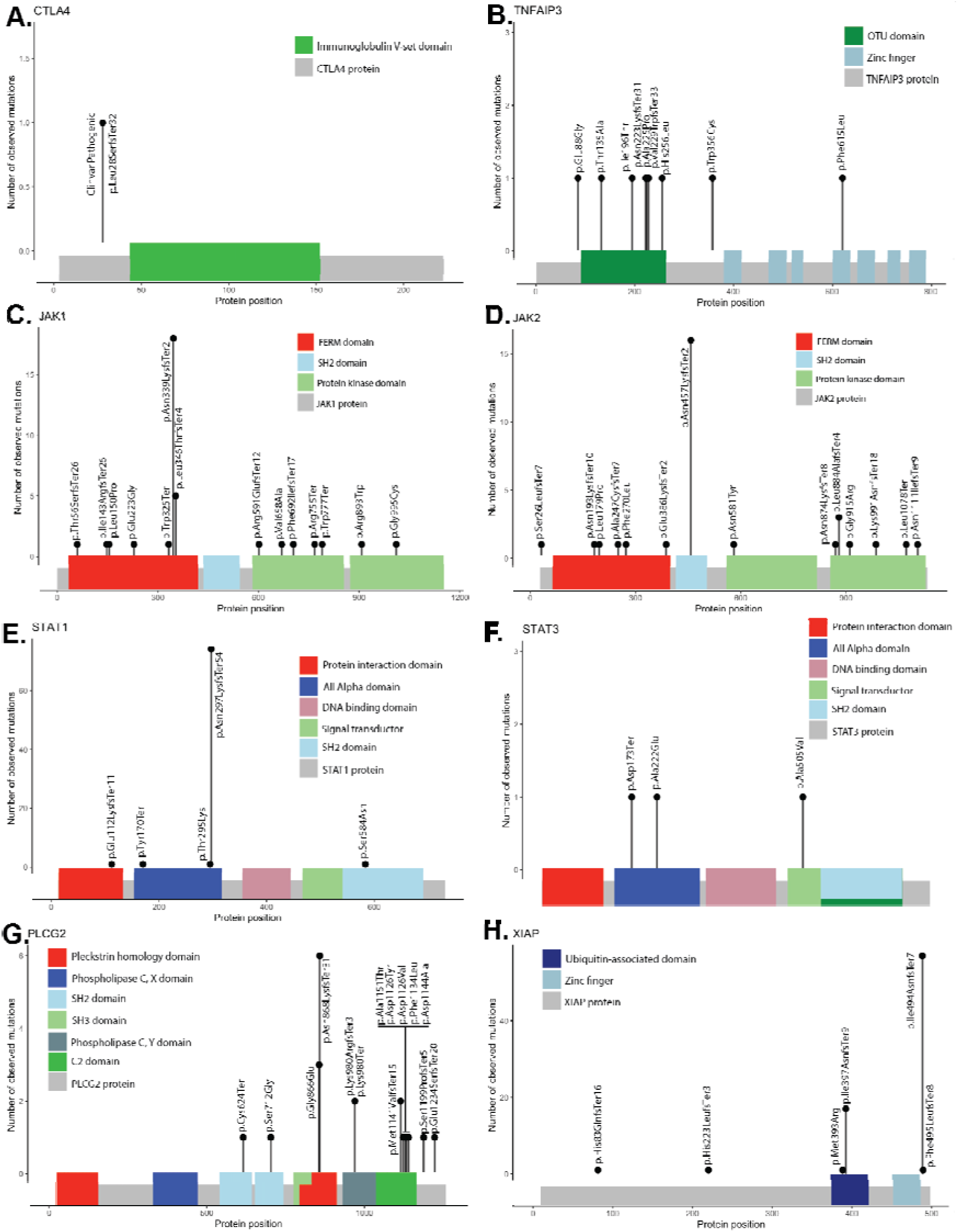
Focused investigation of genes of interest. **A–H**) Somatic P/LP mutations shown for A) CTLA4, B) TNFAIP, C) JAK1, D) JAK2, E) STAT1, F) STAT3, G) PLCG2 and H) XIAP. Bar height indicates the number of times each mutation was observed in the overall cohort. Each mutation is annotated with its effect on the protein sequence. X-axis indicates protein size and mutation location. Important domains in each protein are highlighted for interpretation of the potential effect of each mutation. One highlighted mutation in CTLA4 was previously reported in Clinvar.

We identified 147 pLP-P mutations within four genes of the JAK-STAT pathway: *JAK1*, *JAK2*, *STAT1* and *STAT3*. In *JAK1*, we observed 35 LP somatic mutations across the whole gene over multiple biopsies. We also observed a peak of the same mutation in 18 different biopsies from 17 different patients, within the FERM domain responsible for the receptor interaction. *JAK2*, with 31 LP somatic mutations, showed a similar spread of mutations across important domains, but with a peak in 16 biopsies from 16 patients containing the same mutation in the SH2 domain. In *STAT1*, we observed a pLP-P mutation in the all-alpha transcription factor domain in 74 biopsies, in addition to four other mutations across the gene. In *STAT3*, we observed three somatic mutations: two in the all-alpha domain and one in the signal transductor domain.

We also focused on four genes not in the JAK-STAT pathway: *CTLA4*, *PLCG2*, *XIAP* and *TNFAIP3*.

We observed a frameshift insertion in the beginning of *CTLA4* gene that is annotated as P in Clinvar. This mutation leads to autoimmune lymphoproliferative syndrome due to *CTLA4* haploinsufficiency[30]. We also found 21 pLP-P mutations across many important domains in *PLCG2*. One mutation is in the critical SH2 autoinhibitory domain, and we observed the same somatic mutation six times in the pleckstrin homology domain responsible for transporting the protein to the correct membrane. Five different mutations were observed in the C2 domain where gain-of-function mutations have been described as causal for autoimmune disorders[31]. We also observed 78 pLP-P mutations across *XIAP*, a gene previously observed to cause inflammation in the presence of commensal bacteria when it is contains specific mutations[32], out of which 18 mutations in the ubiquitin-associated domain. Finally, we report nine pLP-P mutations in *TNFAIP3*, many of which cluster in the functional ovarian tumor domain[33]. Germline mutations in *TNFAIP3* can lead to CD-like symptoms in the intestine[34].

### Disease linked pathogenic mutations are observed in intestinal biopsies of IBD patients

We observed 273 pathogenic mutations in the PID, VEO-IBD and GWAS Candidate gene sets that were previously reported in Clinvar. When only considering mutations that cause disease when inherited in an autosomal dominant or X-linked (in males only) manner, we observe pathogenic somatic mutations in four VEO-IBD genes: *CTLA4*, *GUCY2C*, *STAT1* and *IL2RG*. Only the c.2270dup frameshift in *GUCY2C* was observed in more than one IBD patient (**Table 2A**), while 26 different Clinvar-reported pathogenic mutations were observed in PID genes (**Table 2B**) and five were observed in GWAS Candidate genes (**Table 2C**). We observed many more non-autosomal dominant or X-linked mutations in these gene panels (**Supplementary tables S8–S10**).

**Table 2.**
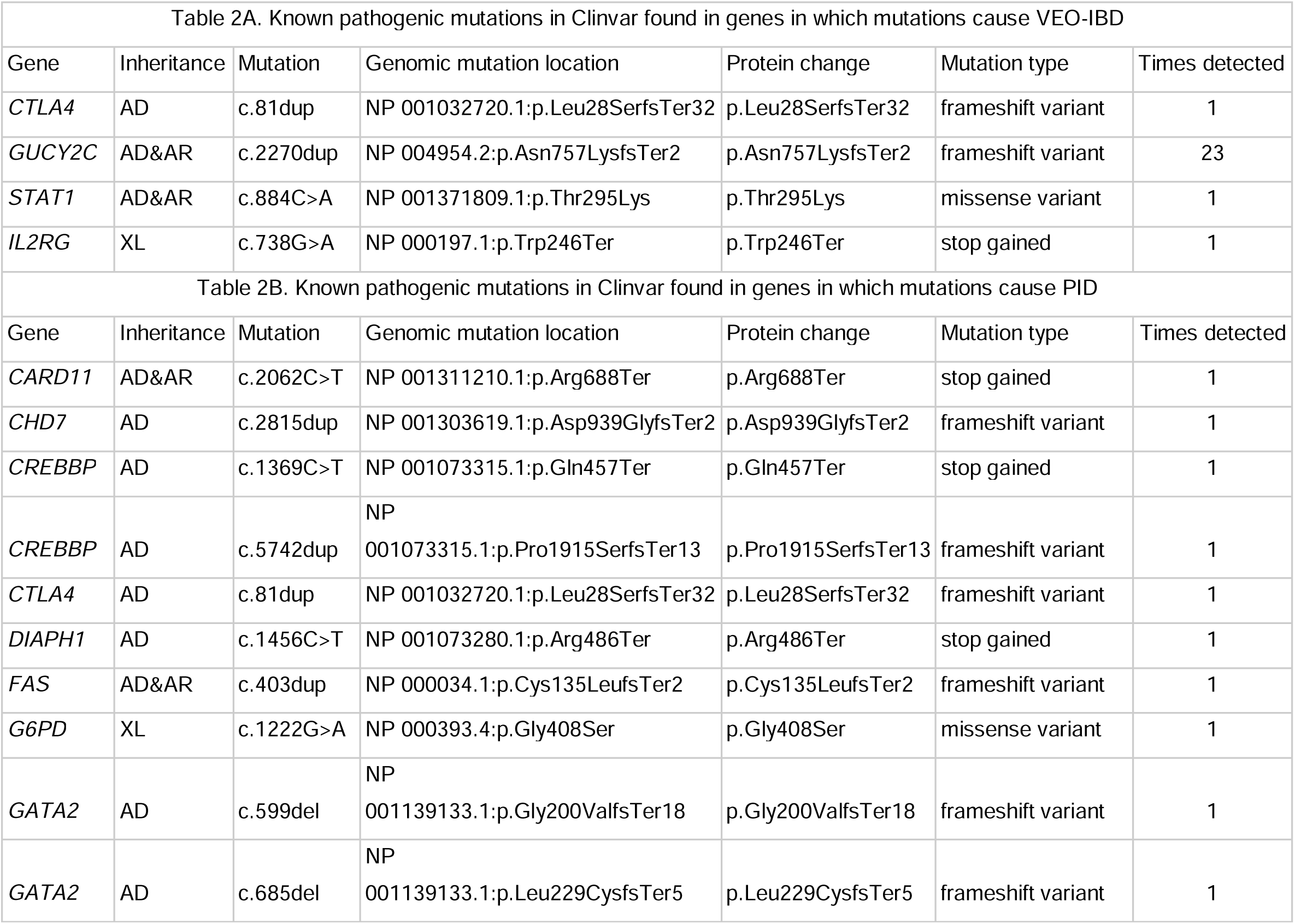
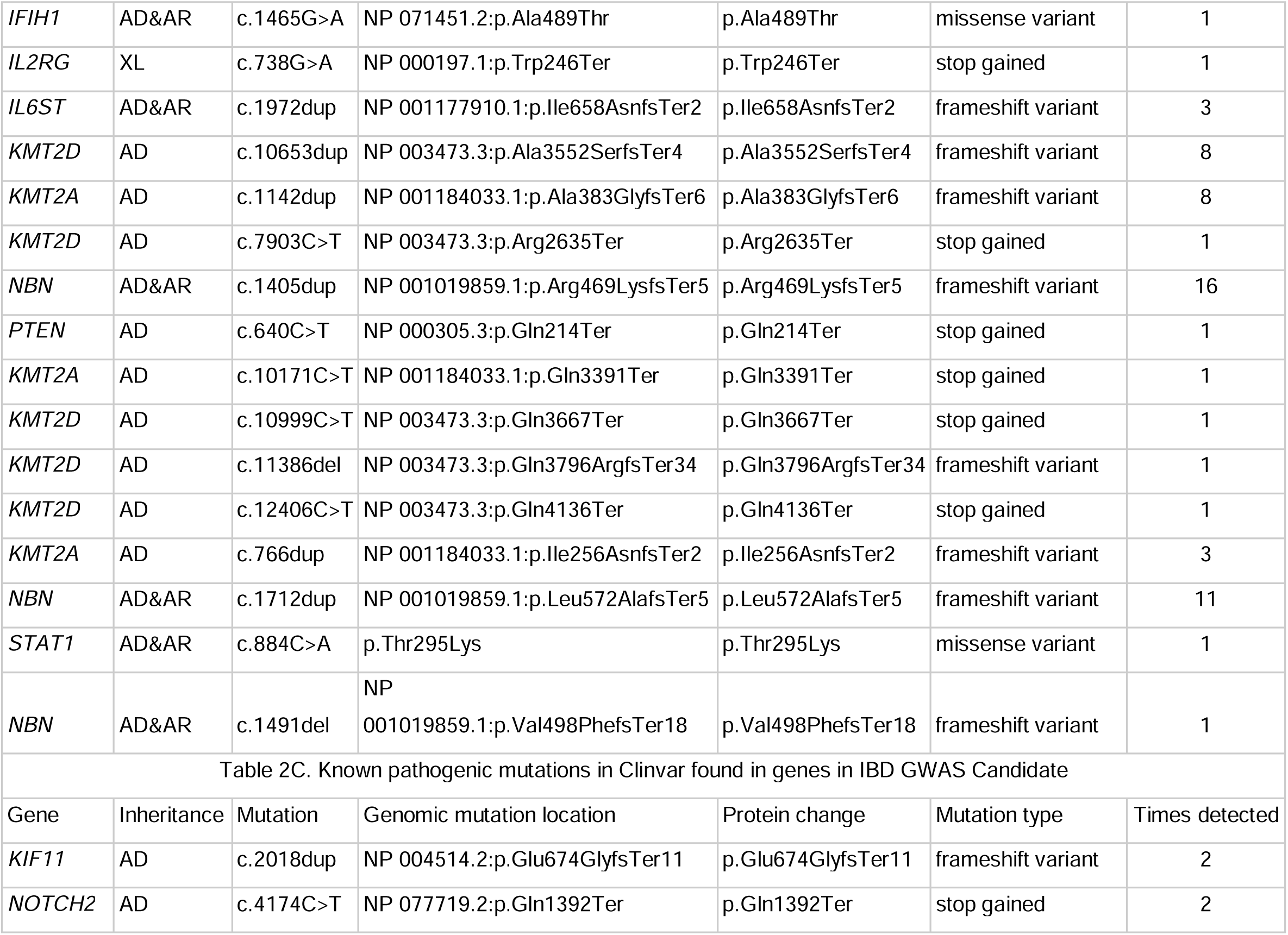
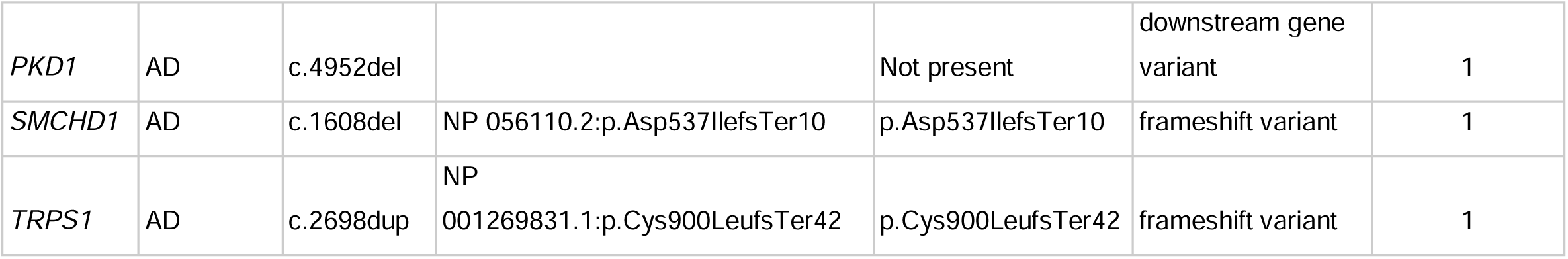
Clinvar annotated Pathogenic mutations in VEO-IBD, PID, and GWAS Candidate genes. Inheritance Autosomal dominant (AD), autosomal recessive (AR) or X-linked (XL) (males only)

## Discussion

We mapped the somatic mutation landscape of the IBD-affected gut using a large patient cohort. Our results reveal large, widespread accumulation of potentially disease-causing mutations in IBD-related genes in IBD patients but not in non-IBD controls. Some of these somatic mutations are in monogenic VEO-IBD and PID genes that can cause intestinal inflammation. For example, the stop gain mutation in *IL2RG* and the frameshift mutation in *STAT1* we observed have a clear relation with IBD pathogenesis as inflammation signaling genes[35,36]. We also report 273 pathogenic somatic mutations in IBD-related genes previously reported in the Clinvar database. These mutations have been confirmed to cause disease when present in the germline.

### Other evidence for accumulation of somatic mutations

Several previous studies have implicated *NFKBIZ*, *ZC3H12A*, *FBXW7*, *ARID1A* and *PIGR* [8,9,37] as genes in which somatic mutations accumulate in IBD. *NFKBIZ* is in our GWAS Candidate gene panel, and we identified seven pLP-P mutations in this gene, revealing a small but overlapping signal. We also observe six pLP-P mutations in *ZC3H12A*, 14 in *FBXW7* and 17 in *ARID1A*. Notably, we did not find any pLP-P mutations in *PIGR.* An overview of these mutations is shown in **Supplementary table S36**. Overall, we have shown reproducible signals in a larger cohort than previously studied, establishing consistent signals between IBD patient groups. We do not observe a relationship between age or disease duration and the number of somatic mutations identified. While this relation has been reported before [8], the correlation between disease duration and somatic mutations is not always present and the effect size of the relationship varies [9].

### Using RNAseq to call somatic mutations: advantages and limitations

In this study we called somatic mutations from RNAseq data, as has been successfully done previously [38]. Calling mutations from expression data has two major advantages. First, we are sure that any mutation that passes our filters is expressed in the relevant tissue, so it is likely to have biological impact. Second, compared to studies that isolate intestinal crypts [8,9,37], we can detect mutations in a wide range of cell types, including recruited immune cells [39]. However, calling mutations from RNAseq data also has limitations. Most other studies have focused on calling somatic mutations from DNA, making it difficult to directly compare observed or expected rates of mutations. Additionally, RNA is under the influence of RNA-editing events [22,23], which must be corrected for. RNAseq data can also be prone to artifacts [40]. To address these limitations, we applied strict filters based on sequencing quality, read depth and VAF and other RNAseq-specific filters [20,24]. While these filters removed many artifacts, they probably also removed some true somatic mutation results. Because of the strict filtering requirement, we are unable to detect mutations in genes with low expression. We were also less likely to detect mutations that reduce gene expression or influence the stability of the RNA molecule [41].

### Clinical implications of somatic mutations in IBD

We found somatic mutations accumulating in IBD-treatment-related pathways, which could lead to possible treatment targets for patients. We showed examples of mutations in the JAK-STAT pathway, known to affect cell signaling in inflammation [42]. However this pathway is highly complex, and both gain- and loss-of-function mutations can have a major impact on prolonging or preventing inflammation [43]. Several genes within this pathway can cause monogenic IBD, inherited in an autosomal dominant manner, meaning a single mutation in one allele is enough to cause disease. Furthermore, there are JAK-inhibiting drugs available on the market, such as tofacitinib, filgotinib and upadacitinib. Potentially, after identifying an accumulation of somatic mutations within this pathway in a patient, one might envision prioritizing this class of drugs to help alleviate this inflammation. Similarly, we could identify different pathways (IL-23/IL-12 pathway, Thiopurine pathway) as potential drug targets through identification of somatic mutations. As we cannot check the size of the area that these mutations spread to, it is difficult to ascertain the specific effect of any of the identified mutations on the etiology or pathogenesis of IBD. However, it has previously been shown that these mutations are able to spread over large areas and therefore have the ability to affect significant amounts of tissue [9,37]. We believe that the combined accumulation of pLP-P somatic mutations we found in IBD-related genes plays an important role in IBD, and this additional genetic effect should be considered in future studies attempting to unravel the complexity of IBD, starting by functionally validating the potential pathogenic effect of these mutations on the intestinal tissue.

## Conclusion

We have uncovered expressed somatic mutations in the intestine of patients with IBD that are enriched in genes strongly associated with IBD or IBD-like symptoms. We identify that IBD genetics goes beyond GWAS associations and risk SNPs, revealing an inflammation-driven dynamic genetic effect potentially leading to an evolving disease. By doing so, we shed light on a previously underexplored area of IBD etiology and pathogenesis.

## Supporting information

Supplemental overview

## Data Availability

https://ega-archive.org/studies/EGAS00001002702

## Authors contribution

FI, AVV and RKW designed the study. IH performed the statistical analysis and drafted the manuscript. BHJ handled the samples in the laboratory. WTKM, GBdV and MAS were involved in the development of the Variant Interpretation Pipeline. MvG was involved in the variant analysis strategy and interpretation. HMvD, MCV, GD, EAMF and RKW included patients. ARB provided support on clinical details. SH, SAS and RAAAR were involved in the project discussions. All authors critically reviewed the manuscript.

## Patient and public involvement

Patients were not involved in the design, conduct, reporting or dissemination plans of this research.

## References

1 Kumar A, Yassin N, Marley A, et al. Crossing barriers: the burden of inflammatory bowel disease across Western Europe. Ther Adv Gastroenterol. 2023;16:17562848231218615. doi: 10.1177/17562848231218615

2 Jostins L, Ripke S, Weersma RK, et al. Host-microbe interactions have shaped the genetic architecture of inflammatory bowel disease. Nature. 2012;491:119–24. doi: 10.1038/nature11582

3 Imhann F, Vich Vila A, Bonder MJ, et al. Interplay of host genetics and gut microbiota underlying the onset and clinical presentation of inflammatory bowel disease. Gut. 2018;67:108–19. doi: 10.1136/gutjnl-2016-312135

4 van der Sloot KWJ, Weersma RK, Alizadeh BZ, et al. Identification of Environmental Risk Factors Associated With the Development of Inflammatory Bowel Disease. J Crohns Colitis. 2020;14:1662–71. doi: 10.1093/ecco-jcc/jjaa114

5 Vich Vila A, Imhann F, Collij V, et al. Gut microbiota composition and functional changes in inflammatory bowel disease and irritable bowel syndrome. Sci Transl Med. 2018;10:eaap8914. doi: 10.1126/scitranslmed.aap8914

6 Liu Z, Liu R, Gao H, et al. Genetic architecture of the inflammatory bowel diseases across East Asian and European ancestries. Nat Genet. 2023;55:796–806. doi: 10.1038/s41588-023-01384-0

7 Sazonovs A, Stevens CR, Venkataraman GR, et al. Large-scale sequencing identifies multiple genes and rare variants associated with Crohn’s disease susceptibility. Nat Genet. 2022;54:1275–83. doi: 10.1038/s41588-022-01156-2

8 Olafsson S, McIntyre RE, Coorens T, et al. Somatic Evolution in Non-neoplastic IBD-Affected Colon. Cell. 2020;182:672–684.e11. doi: 10.1016/j.cell.2020.06.036

9 Nanki K, Fujii M, Shimokawa M, et al. Somatic inflammatory gene mutations in human ulcerative colitis epithelium. Nature. 2020;577:254–9. doi: 10.1038/s41586-019-1844-5

10 Luzzatto L. Somatic mutations in cancer development. Environ Health. 2011;10:S12. doi: 10.1186/1476-069X-10-S1-S12

11 Zaidi SH, Harrison TA, Phipps AI, et al. Landscape of somatic single nucleotide variants and indels in colorectal cancer and impact on survival. Nat Commun. 2020;11:3644. doi: 10.1038/s41467-020-17386-z

12 Lei H, Tao K. Somatic mutations in colorectal cancer are associated with the epigenetic modifications. J Cell Mol Med. 2020;24:11828–36. doi: 10.1111/jcmm.15799

13 Alexandrov LB, Nik-Zainal S, Wedge DC, et al. Signatures of mutational processes in human cancer. Nature. 2013;500:415–21. doi: 10.1038/nature12477

14 Terlouw D, Suerink M, Boot A, et al. Recurrent APC Splice Variant c.835-8A>G in Patients With Unexplained Colorectal Polyposis Fulfilling the Colibactin Mutational Signature. Gastroenterology. 2020;159:1612–1614.e5. doi: 10.1053/j.gastro.2020.06.055

15 Pleguezuelos-Manzano C, Puschhof J, Rosendahl Huber A, et al. Mutational signature in colorectal cancer caused by genotoxic pks+ E. coli. Nature. 2020;580:269–73. doi: 10.1038/s41586-020-2080-8

16 Imhann F, Van der Velde KJ, Barbieri R, et al. The 1000IBD project: multi-omics data of 1000 inflammatory bowel disease patients; data release 1. BMC Gastroenterol. 2019;19:5. doi: 10.1186/s12876-018-0917-5

17 van der Velde KJ, Imhann F, Charbon B, et al. MOLGENIS research: advanced bioinformatics data software for non-bioinformaticians. Bioinforma Oxf Engl. 2019;35:1076–8. doi: 10.1093/bioinformatics/bty742

18 Hu S, Uniken Venema WT, Westra H-J, et al. Inflammation status modulates the effect of host genetic variation on intestinal gene expression in inflammatory bowel disease. Nat Commun. 2021;12:1122. doi: 10.1038/s41467-021-21458-z

19 Hu S, Bourgonje AR, Gacesa R, et al. Mucosal host-microbe interactions associate with clinical phenotypes in inflammatory bowel disease. Nat Commun. 2024;15:1470. doi: 10.1038/s41467-024-45855-2

20 Van der Auwera GA, Carneiro MO, Hartl C, et al. From FastQ data to high confidence variant calls: the Genome Analysis Toolkit best practices pipeline. Curr Protoc Bioinforma Ed Board Andreas Baxevanis Al. 2013;11:11.10.1–11.10.33. doi: 10.1002/0471250953.bi1110s43

21 Dobin A, Davis CA, Schlesinger F, et al. STAR: ultrafast universal RNA-seq aligner. Bioinformatics. 2013;29:15–21. doi: 10.1093/bioinformatics/bts635

22 Ramaswami G, Li JB. RADAR: a rigorously annotated database of A-to-I RNA editing. Nucleic Acids Res. 2014;42:D109–13. doi: 10.1093/nar/gkt996

23 Kiran A, Baranov PV. DARNED: a DAtabase of RNa EDiting in humans. Bioinforma Oxf Engl. 2010;26:1772–6. doi: 10.1093/bioinformatics/btq285

24 García-Nieto PE, Morrison AJ, Fraser HB. The somatic mutation landscape of the human body. Genome Biol. 2019;20:298. doi: 10.1186/s13059-019-1919-5

25 Maassen WTK, Johansson LF, Charbon B, et al. MOLGENIS VIP: an open-source and modular pipeline for high-throughput and integrated DNA variant analysis. 2024;2024.04.11.24305656.

26 Li S, van der Velde KJ, de Ridder D, et al. CAPICE: a computational method for Consequence-Agnostic Pathogenicity Interpretation of Clinical Exome variations. Genome Med. 2020;12:75. doi: 10.1186/s13073-020-00775-w

27 Richards S, Aziz N, Bale S, et al. Standards and Guidelines for the Interpretation of Sequence Variants: A Joint Consensus Recommendation of the American College of Medical Genetics and Genomics and the Association for Molecular Pathology. Genet Med Off J Am Coll Med Genet. 2015;17:405–24. doi: 10.1038/gim.2015.30

28 Karczewski KJ, Francioli LC, Tiao G, et al. The mutational constraint spectrum quantified from variation in 141,456 humans. Nature. 2020;581:434–43. doi: 10.1038/s41586-020-2308-7

29 Soheili-Nezhad S, van der Linden RJ, Olde Rikkert M, et al. Long genes are more frequently affected by somatic mutations and show reduced expression in Alzheimer’s disease: Implications for disease etiology. Alzheimers Dement. 2021;17:489–99. doi: 10.1002/alz.12211

30 Collen LV, Salgado CA, Bao B, et al. Cytotoxic T Lymphocyte Antigen 4 Haploinsufficiency Presenting As Refractory Celiac-Like Disease: Case Report. Front Immunol. 2022;13:894648. doi: 10.3389/fimmu.2022.894648

31 Novice T, Kariminia A, Del Bel KL, et al. A Germline Mutation in the C2 Domain of PLCγ2 Associated with Gain-of-Function Expands the Phenotype for PLCG2-Related Diseases. J Clin Immunol. 2020;40:267–76. doi: 10.1007/s10875-019-00731-3

32 Wahida A, Müller M, Hiergeist A, et al. XIAP restrains TNF-driven intestinal inflammation and dysbiosis by promoting innate immune responses of Paneth and dendritic cells. Sci Immunol. 2021;6:eabf7235. doi: 10.1126/sciimmunol.abf7235

33 Chen Y, Ye Z, Chen L, et al. Association of Clinical Phenotypes in Haploinsufficiency A20 (HA20) With Disrupted Domains of A20. Front Immunol. 2020;11:574992. doi: 10.3389/fimmu.2020.574992

34 Yazısız V. Similarities and differences between Behçet’s disease and Crohn’s disease. World J Gastrointest Pathophysiol. 2014;5:228–38. doi: 10.4291/wjgp.v5.i3.228

35 Schreiber S, Rosenstiel P, Hampe J, et al. Activation of signal transducer and activator of transcription (STAT) 1 in human chronic inflammatory bowel disease. Gut. 2002;51:379–85.

36 Ogawa A, Watanabe T, Natsume T, et al. Early-Onset Inflammatory Bowel Disease Caused by Mutations in the X-Linked Gene IL2RG. J Investig Allergol Clin Immunol. 2021;31:69–71. doi: 10.18176/jiaci.0523

37 Kakiuchi N, Yoshida K, Uchino M, et al. Frequent mutations that converge on the NFKBIZ pathway in ulcerative colitis. Nature. 2020;577:260–5. doi: 10.1038/s41586-019-1856-1

38 Yizhak K, Aguet F, Kim J, et al. RNA sequence analysis reveals macroscopic somatic clonal expansion across normal tissues. Science. 2019;364:eaaw0726. doi: 10.1126/science.aaw0726

39 Saez A, Herrero-Fernandez B, Gomez-Bris R, et al. Pathophysiology of Inflammatory Bowel Disease: Innate Immune System. Int J Mol Sci. 2023;24:1526. doi: 10.3390/ijms24021526

40 Verwilt J, Mestdagh P, Vandesompele J. Artifacts and biases of the reverse transcription reaction in RNA sequencing. RNA. 2023;29:889–97. doi: 10.1261/rna.079623.123

41 Agarwal V, Kelley DR. The genetic and biochemical determinants of mRNA degradation rates in mammals. Genome Biol. 2022;23:245. doi: 10.1186/s13059-022-02811-x

42 Coskun M, Salem M, Pedersen J, et al. Involvement of JAK/STAT signaling in the pathogenesis of inflammatory bowel disease. Pharmacol Res. 2013;76:1–8. doi: 10.1016/j.phrs.2013.06.007

43 Ott N, Faletti L, Heeg M, et al. JAKs and STATs from a Clinical Perspective: Loss-of-Function Mutations, Gain-of-Function Mutations, and Their Multidimensional Consequences. J Clin Immunol. 2023;43:1326–59. doi: 10.1007/s10875-023-01483-x

