## Supplemental overview for "Intestinal somatic mutations in inflammatory bowel disease patients are enriched in very early onset IBD and primary immunodeficiency genes"

and

Prof. Dr. Rinse K. Weersma, Department of Gastroenterology and Hepatology, University of Groningen and University Medical Center Groningen, P.O. Box 30.001, Groningen 9700 RB, the Netherlands;

### Index Supplementary Appendix

**Supplementary Appendix. Online methods**

**Supplementary Table S1. Sample description**

**Supplementary Table S2. VEO-IBD gene list**

**Supplementary Table S3. PID gene list**

**Supplementary Table S4. GWAS Candidate gene list**

**Supplementary Table S5. GWAS Loci gene list**

**Supplementary Table S6. Database Annotated mutation overview**

**Supplementary Table S7. Pathogenic/likely pathogenic annotation inheritance overview**

**Supplementary Table S8. Clinvar VEO-IBD non-AD mutations**

**Supplementary Table S9. Clinvar PID non-AD mutations**

**Supplementary Table S10. Clinvar GWAS Candidate non-AD mutations**

**Supplementary table S11. All VKGL mutations**

**Supplementary table S12. All Clinvar mutations**

**Supplementary table S13. VEO-IBD VKGL**

**Supplementary table S14. VEO-IBD Clinvar**

**Supplementary table S15. GWAS Candidate VKGL**

**Supplementary table S16. GWAS Candidate Clinvar**

**Supplementary table S17. GWAS Loci VKGL**

**Supplementary table S18. GWAS Loci Clinvar**

**Supplementary table S19. PID VKGL**

**Supplementary table S20. PID Clinvar**

**Supplementary table S21. GWAS Candidate all LP**

**Supplementary table S22. GWAS Candidate AD**

**Supplementary table S23. GWAS Candidate AR**

**Supplementary table S24. GWAS Loci All LP**

**Supplementary table S25. GWAS Loci AD**

**Supplementary table S26. GWAS Loci AR**

**Supplementary table S27. GWAS Loci X-linked males**

**Supplementary table S28. PID All LP**

**Supplementary table S29. PID AD**

**Supplementary table S30. PID AR**

**Supplementary table S31. PID X-linked males**

**Supplementary table S32. VEO-IBD All LP**

**Supplementary table S33. VEO-IBD AD**

**Supplementary table S34. VEO-IBD AR**

**Supplementary table S35. VEO-IBD X-linked males**

**Supplementary table S36. Mutations in previously identified genes**

**Supplementary Figure S1. Somatic mutation distribution**

**Supplementary Figure S2. Somatic mutations over age at biopsy**

**Supplementary Figure S3. P/LP somatic mutations in cancer genes over age at biopsy**

**Supplementary Figure S4. Somatic mutations over inflammation status**

**Supplementary Figure S5. P/LP somatic mutations over inflammation status**

#### Supplementary Methods

##### Cohort description

In this study, we used data generated from intestinal biopsies and blood samples collected at the University Medical Center Groningen (UMCG). In total, 268 participants with a confirmed inflammatory bowel disease (IBD) diagnosis (Crohn's disease (CD) or ulcerative colitis (UC)) and 17 non-IBD control participants were included. Of the 268 IBD participants, 58% were female and 55% were diagnosed with CD (see Table 1 for a full overview). Participants were at least 18 years old and were enrolled between 2003 and 2019. Diagnosis of IBD was based on clinical, laboratory, endoscopic and histopathological investigation. Detailed phenotypic data, including age, sex, BMI, smoking status, Montreal disease classification, medication usage, history of surgery and clinical disease activity were collected for all patients, at the time of inclusion. Montreal disease classification was recorded from the closest visit to the outpatient clinic at the time of sampling. Clinical disease activity was established using the Harvey-Bradshaw Index for patients with CD and with the Simple Clinical Colitis Activity Index for patients with UC. Prior to sample collection, all participants provided written informed consent.

##### RNA

RNA was isolated from these biopsies using the AllPrep DNA/RNA mini kit from Qiagen, according to the manufacturer's instructions. The intestinal biopsies were homogenized using RLT lysis buffer with Beta-mercaptoethanol using the Qiagen TissueLyser with stainless steel beads. RNA-sequencing was performed in two batches. For the first batch (147 samples), sample preparation was done using the BioScientific NEXTflex Rapid Directional RNA-Seq kit (Perkin-Elmer), followed by paired-end sequencing of RNA using Illumina NextSeq500 sequencing. For the second batch (442 samples), sample preparation was performed for construction of the Eukaryotic Transcriptome Library (Novogene), followed by paired-end sequencing using the Illumina HiSeq PE250 platform.

##### WES

Blood plasma samples were available for the 268 IBD participants. All were collected at a clinical visit prior to the colonoscopy sample collection. DNA isolation was done with the AutoPure LS procedure from Qiagen. The WES data was generated at the Broad Institute of Harvard and MIT (Boston, USA), using the Illumina HiSeq 2500 sequencing platform. After quality control with default parameters from the GATK best practices, the reads were then aligned to the hg38 human reference genome.

**Prediction of somatic mutations**

As no WES data were available for the control samples, we used GATK Mutect2 in tumor-only mode on both control and IBD biopsies. For this purpose, we used the GATK best practices for variant discovery. The RNAseq data from all biopsies was aligned using STAR aligner (v2.7.3a) with the outSAMmapqUnique option set to 60, as recommended by GATK best practices. Duplicate reads were then marked using MarkDuplicatesSpark (GATK v4.2.4.1) to prevent artifacts from being annotated as real mutations, and base quality scores were recalculated using BaseRecalibrator (GATK v4.2.4.1). These bam files were then used as input for the GATK Mutect2 tool in tumor-only mode, using a panel of normals from GATK and hg38 as reference genome. Additionally, gnomAD data was included as a germline resource using the af-only-gnomad.hg38.vcf.gz file provided by GATK. Afterwards, Funcotator (GATK v4.2.4.1) was used to annotate the mutations.

**Identification of somatic mutations**

To identify somatic mutations in the intestinal tract of participants with IBD, we combined germline genotypes obtained from WES data with RNAseq gene expression of intestinal biopsies. To achieve this, GATK best practices for variant discovery were followed. The RNAseq reads were aligned to the hg38 reference genome using STAR aligner (v2.7.3a) with the outSAMmapqUnique option set to 60, as recommended by GATK best practices. Duplicate reads were then marked using MarkDuplicatesSpark (GATK v4.2.4.1) to prevent artifacts from being annotated as real mutations, and base quality scores were recalculated using BaseRecalibrator (GATK v4.2.4.1). The matched RNAseq and WES data for each participant was subsequently used as input for GATK mutect2. GATK Mutect2 was used with the dont-use-soft-clipped-bases setting as True. A panel of normals based on hg38 was included in the run, and hg38 was used as the reference genome. Additionally, gnomAD data was included as a germline resource using the af-only-gnomad.hg38.vcf.gz file provided by GATK. Afterwards, Funcotator (GATK v4.2.4.1) was used to annotate the mutations.

**Quality control on predicted somatic mutations.**

Many sources of artifacts or other errors must be considered when calling somatic mutations from RNAseq data. For this purpose, we used multiple filtering steps, including some taken from the pipeline created by Garcia-Nieto et al[24]. First, the FilterMutectCalls function from GATK was used to filter low confidence mutation predictions. On average, this step filtered out about 60% of the initial predicted mutations. Next, a filter was applied based on minimum coverage and a minimum percentage of reads supporting the alternative allele to increase the confidence of the mutation predictions. Estimated mutations were required to have a minimum of 10 reads on the location of each mutation, with at least 10% or 25% of the reads supporting the alternative allele. This step filtered out 74% and 90% of the remaining predicted mutations, respectively. The results of both filters were reported, but only mutations called with the 25% filter were considered for functional interpretation. Finally, the mutations were filtered on splice junctions and RNA-editing sites. Splice junction mutations were filtered out because an increase in mutation calls from RNA was previously observed close to exon boundaries that was not observed in the DNA. Mutations in RNA-editing sites were filtered out because this process introduces mutations after RNA transcription, and these are not somatic mutations. We therefore removed any mutation called in known RNA-editing sites from the RADAR and DARNED databases. These databases were downloaded for the pipeline, and the UCSC liftover tool was used to convert the hg19 versions of the databases to the hg38 version. Filtering on splice junctions removed 20% of the remaining predicted mutations, and filtering on RNA-editing removed another 5%. For the interpretation of deleteriousness, we removed somatic mutations labeled by Funcotator as predicted in RNA and pseudogenes, which accounted for 8% of the remaining predicted mutations. In total, an average of 29,611 predicted mutations per biopsy were filtered out, leaving an average of 743 mutations per biopsy for the downstream analysis.

**Determination of genes without somatic mutations**

To determine which genes in the analysis had no mutations, we determined the coverage for the RNAseq and the WES data over all genes in the WES gene panel. For this purpose, the aligned bam files were used as input for the bedtools (v2.30.0) coverage function and overlapped with Refseq exon locations for hg38. A cut-off of at least 10 reads covering each gene was used. This minimum, combined with our minimum coverage cut-off for mutation-calling, ensured an accurate determination of the number of mutations per gene, including zero or lack of detection.

**Somatic mutations related to age or cancer genes**

To analyze the relationship between age and somatic mutations, we created a linear model comparing age and the number of somatic mutations, both for all mutations and after P/LP annotation. Additionally, to investigate the accumulation of somatic mutations in genes associated with cancer, we considered genes from the National Cancer Institute with a mutation in cancer in at least 20% of their cohort, based on the colorectal cancer set and mutation frequency dataset with cancer gene census turned off (last check 19 March 2024). This gave the following gene list: *APC*, *TP53*, *TTN*, *KRAS*, *MUC16*, *SYNE1*, *FAT4*, *PIK3CA*, *OBSCN*, *ZFHX4*, *RYR2* and *CSMD3*. For these genes, we also created linear models relating age and the number of mutations in these genes for both all mutations and only P/LP mutations. We also added all the mutations in these genes together and investigated the linear relationship between this cumulative cancer mutation load and age.

**Statistical analysis**

The R package lme4 (v1.1.33) was used to model statistical differences between the number of P/LP somatic mutations per gene set over different phenotypes using linear mixed models. Initially, we tested each phenotype alone in a univariable model only correcting for within-patient effects.

To determine the difference in somatic mutations between controls and IBD we designed a model correcting for age, sex, BMI, location and inflammation status of the biopsy as well correcting for multiple samples per patient. This was done for all mutations as well at the number of mutations per gene set, as described by the following models:

Overall Number of mutations = Intercept + Diagnosis + Age + Sex + Location of biopsy + inflammation status+ 1 | Patient id

Number of mutations in VEO-IBD genes = Intercept + Diagnosis + Age + Sex + Location of biopsy + inflammation status+ 1 | Patient id

Number of mutations in GWAS-candidate genes = Intercept + Diagnosis + Age + Sex + Location of biopsy + inflammation status+ 1 | Patient id

Number of mutations in GWAS-loci genes = Intercept + Diagnosis + Age + Sex + Location of biopsy + inflammation status+ 1 | Patient id

Number of mutations in PID genes = Intercept + Diagnosis + Age + Sex + Location of biopsy + inflammation status+ 1 | Patient id

We then created a model to test the effect of inflammation per gene set, correcting for sequencing batch, biopsy tissue location and a within-patient effect.

Our inflammation model is described by the following formula:

Inflammation = Intercept + Number of mutations in gene set + Location of biopsy + Sequencing batch + 1 | Patient id

The output of these models was shown using the Emeans (v1.8.6) package.

The numerical differences in number of annotated mutations per gene sets were determined using chi-squared tests. And the comparison within IBD biopsies was done with a Wilcoxon signed-rank also known as paired sample Mann-Whitney test.

**Supplementary table S1. Biopsies**

| | IBD Inflamed (n=198) | IBD non-inflamed (n=340) | Non-IBD control (n=51) | P-value between inflamed and non-inflamed |
|---|---|---|---|---|
| **Location, no. (%)** | | | | |
| Colon | 148 (74.7) | 208 (61.2) | 42 (82.4) | 0.00184 |
| Ileum | 50 (25.3) | 132 (38.8) | 9 (17.6) | 0.0018 |
| **Diagnosis, no. (%)** | | | | |
| CD | 105 (53.0) | 201 (59.1) | 0 (0) | 0.199 |
| UC | 93 (47.0) | 139 (40.9) | 0 (0) | |
| **Sex, no. (%)** | | | | |
| Female | 108 (54.5) | 203 (59.7) | 17 (33.3) | 0.281 |
| **Age at biopsy, mean (SD)** | 41.7 (16.1) | 43.2 (16.0) | 45.9 (11.0) | 0.306 |
| **BMI, mean (SD)** | 25.1 (4.55) | 25.2 (4.71) | 24.6 (3.3) | 0.885 |
| **Smoking, yes (%)** | 39 (19.7) | 73 (21.5) | 20 (39.2) | 0.679 |
| **Medication, yes (%)** | | | | |
| Mesalazines | 89 (44.9) | 138 (40.6) | 0 (0) | 0.37 |
| Steroids | 117 (59.1) | 123 (36.2) | 0 (0) | 4.05E-07 |
| Thiopurines | 59 (29.8) | 116 (34.1) | 0 (0) | 0.35 |
| Methotrexate | 19 (9.6) | 24 (7.1) | 0 (0) | 0.38 |
| Anti-TNF | 24 (12.1) | 49 (14.4) | 2 (4) | 0.54 |
| **Montreal classification, no. (%)** | | | | |
| **Montreal A (1 missing)** | | | | |
| A1: <17 years | 22 (11.1) | 45 (13.2) | NA | |
| A2: 17–40 years | 122 (61.6) | 197 (57.9) | NA | |
| A3: >40 years | 54 (27.3) | 97 (28.5) | NA | 0.668 |
| **Within CD** | | | | |
| **Montreal L (11 missing)** | | | | |
| L1 (+L4) | 11 (+0) | 49 (+6) | NA | 0.016 |

| | | | | |
|---|---|---|---|---|
| L2 (+L4) | 19 (+1) | 34 (+2) | NA | |
| L3 (+L4) | 70 (+7) | 112 (+11) | NA | |
| **Montreal B (11 missing)** | | | | |
| B1 (p) | 58 (+12) | 111 (+30) | NA | 0.076 |
| B2 (p) | 36 (+17) | 56 (+23) | NA | |
| B3 (p) | 6 (+0) | 28 (+10) | | |
| **Within UC** | | | | |
| **Montreal E (32 missing)** | | | | |
| E1 | 5 | 8 | | 0.943 |
| E2 | 23 | 35 | | |
| E3 | 54 | 75 | | |
| **Montreal S (46 missing)** | | | | |
| S0 | 2 | 6 | | 0.058 |
| S1 | 7 | 17 | | |
| S2 | 31 | 54 | | |
| S3 | 37 | 32 | | |

**Supplementary table S2**

**VEO-IBD gene list (see Excel file)**

**Supplementary table S3**

**PID gene list (see Excel file)**

**Supplementary table S4**

**GWAS Candidate gene list (see Excel file)**

**Supplementary table S5**

**GWAS Loci gene list (see Excel file)**

**Supplementary table S6. Database annotated mutation overview**

**Table S6. Pathogenic and likely pathogenic mutations overview**

| Database | VEO-IBD genes n=103 | PID genes n=475 | GWAS Candidate genes n=318 | GWAS Loci genes n=3504 |
|---|---|---|---|---|
| **VKGL** | 1 | 21 | 13 | 23 |
| **Clinvar** | 36 | 220 | 18 | 257 |

*The number of mutations in each gene set that overlap with the VKGL or Clinvar Pathogenic/Likely Pathogenic database. Despite having fewer genes, the Very Early Onset-IBD (VEO-IBD) and Primary Immune Deficiency (PID) genes show a significantly higher number of pathogenic that were previously reported in the Clinvar database mutations (chi-squared, p=2.88e-87).*

**Supplementary table S7**

**Pathogenic/likely pathogenic annotation inheritance overview**

| | VEO-IBD genes n=103 | PID genes n=475 | GWAS Candidate genes n=318 | GWAS Loci genes n=3504 |
|---|---|---|---|---|
| **AD** | 57 | 1232 | 371 | 2317 |
| **AR** | 356 | 2076 | 654 | 2651 |
| **XL male** | 50 | 142 | 0 | 87 |
| **XL Female** | 64 | 113 | 0 | 27 |
| **Unknown/ unclear inheritance** | 288 | 2355 | 2063 | 12135 |
| **Total** | 777 | 5840 | 2337 | 17169 |

*The number of Pathogenic/Likely Pathogenic mutations annotated by the Variant Interpretation Pipeline, shown per gene set and split over inheritance. Autosomal dominant (AD) and X-linked (XL) males are most likely affected by the identified mutations. Again, despite the low number of genes in the set, Primary Immune Deficiency (PID) genes show many AD or XL male P/LP mutations in IBD biopsies (chi-squared, p=3.15e-139). Very early onset IBD, VEO-IBD.*

**Supplementary table S8. Clinvar very early onset IBD non-autosomal dominant mutations**

| Gene | Inherit-ance | Mutation | Clinvar | CAPICE score | Genomic mutation location | Protein change | VIP Anno-tation | Mutation Type | Times detected |
|---|---|---|---|---|---|---|---|---|---|
| *HPS1* | AR | c.517C>T | P | 0,93620735 | NP 000186.2:p.Arg173Ter | p.Arg173Ter | LP | stop gained | 1 |
| *HPS4* | AR | c.1132C>T | P/LP | 0,6503983 | NP 001336825.1:p.Gln378Ter | p.Gln378Ter | LP | stop gained | 1 |
| *IL10RA* | AR | c.301C>T | P | 0,10837812 | NP 001549.2:p.Arg101Trp | p.Arg101Trp | LP | missense variant | 1 |
| *ITGB2* | AR | c.533C>T | P | 0,18867305 | NP 000202.3:p.Pro178Leu | p.Pro178Leu | LP | missense variant | 1 |
| *LRBA* | AR | c.2614del | P | 0,99832445 | NP 001186211.2:p.Ser872LeufsTer15 | p.Ser872LeufsTer15 | LP | frameshift variant | 1 |
| *RAG1* | AR | c.705dup | LP | 0,9967925 | NP 000439.2:p.Leu236ThrfsTer6 | p.Leu236ThrfsTer6 | LP | frameshift variant | 2 |
| *ZBTB24* | AR | c.501dup | P | 0,99200296 | NP 001157785.1:p.Val168SerfsTer28 | p.Val168SerfsTer28 | LP | frameshift variant | 3 |

*All non-autosomal dominant or X-linked pathogenic mutations in the VEO-IBD gene set previously reported in the Clinvar database. AR, autosomal recessive. P, pathogenic. LP, likely pathogenic.*

**Supplementary table S9. Clinvar primary immune deficiency non-autosomal dominant mutations**

| Gene | Inherit -ance | Mutation | Clinvar | CAPICE score | Genomic mutation location | Protein change | VIP annot. | Mutation Type | Times detected |
|---|---|---|---|---|---|---|---|---|---|
| *ADA2* | AR | c.548T>C | LP | 0,9609852 | NP 001269154.1:p.Leu183Pro | p.Leu183Pro | LP | missense variant | 1 |
| *AP3B1* | AR | c.1642dup | LP | 0,9819754 | NP 001258698.1:p.Ile548AsnfsTer12 | p.Ile548AsnfsTer12 | LP | frameshift variant | 75 |
| *AP3B1* | AR | c.569G>A | P | 0,9977349 | NP 001258698.1:p.Trp190Ter | p.Trp190Ter | LP | stop gained | 2 |
| *BCL10* | AR | c.466dup | P | 0,99654704 | NP 001307644.1:p.Ser156PhefsTer3 | p.Ser156PhefsTer3 | LP | frameshift variant | 7 |
| *CARMIL2* | AR | c.3493C>T | P | 0,9400393 | NP 001013860.1:p.Arg1165Ter | p.Arg1165Ter | LP | stop gained | 1 |
| *CFTR* | AR | c.2089dup | P | 0,99667656 | NP 000483.3:p.Arg697LysfsTer33 | p.Arg697LysfsTer33 | LP | frameshift variant | 11 |
| *CFTR* | AR | c.2052dup | P | 0,8512588 | NP 000483.3:p.Gln685ThrfsTer4 | p.Gln685ThrfsTer4 | LP | frameshift variant | 13 |
| *CFTR* | AR | c.3773dup | P | 0,8142965 | NP 000483.3:p.Leu1258PhefsTer7 | p.Leu1258PhefsTer7 | LP | frameshift variant | 6 |
| *EPG5* | AR | c.6898dup | P | 0,9945332 | NP 066015.2:p.Met2300AsnfsTer25 | p.Met2300AsnfsTer25 | LP | frameshift variant | 8 |
| *EPG5* | AR | c.5704dup | P | 0,9717254 | NP 066015.2:p.Tyr1902LeufsTer2 | p.Tyr1902LeufsTer2 | LP | frameshift variant | 1 |
| *IL10RA* | AR | c.301C>T | P | 0,10837812 | NP 001549.2:p.Arg101Trp | p.Arg101Trp | LP | missense variant | 1 |
| *IL7R* | AR | c.470del | P | 0,54579556 | NP 002176.2:p.Lys157ArgfsTer4 | p.Lys157ArgfsTer4 | LP | frameshift variant | 10 |
| *RASGRP2* | AR | c.1479dup | LP | 0,65579325 | NP 001092140.1:p.Arg494AlafsTer54 | p.Arg494AlafsTer54 | LP | frameshift variant | 1 |
| *RECQL4* | AR | c.3193C>T | P | 0,99029887 | NP 004251.4:p.Gln1065Ter | p.Gln1065Ter | LP | stop gained | 1 |
| *VPS13B* | AR | c.5737dup | P | 0,8790979 | NP 060360.3:p.Ile1913AsnfsTer7 | p.Ile1913AsnfsTer7 | LP | frameshift variant | 2 |
| *VPS13B* | AR | c.6200dup | LP | 0,98977786 | NP 060360.3:p.Leu2067PhefsTer13 | p.Leu2067PhefsTer13 | LP | frameshift variant | 1 |
| *RAG1* | AR | c.705dup | LP | 0,9967925 | NP 000439.2:p.Leu236ThrfsTer6 | p.Leu236ThrfsTer6 | LP | frameshift variant | 1 |

| | | | | | | | | | |
|---|---|---|---|---|---|---|---|---|---|
| *ITGB2* | AR | c.533C>T | P | 0,18867305 | NP 000202.3:p.Pro178Leu | p.Pro178Leu | LP | missense variant | 1 |
| *LRBA* | AR | c.2614del | P | 0,99832445 | NP 001186211.2:p.Ser872LeufsTer15 | p.Ser872LeufsTer15 | LP | frameshift variant | 1 |
| *VPS13B* | AR | c.5608dup | P | 0,89815784 | NP 060360.3:p.Thr1870AsnfsTer12 | p.Thr1870AsnfsTer12 | LP | frameshift variant | 3 |
| *ZBTB24* | AR | c.501dup | P | 0,99200296 | NP 001157785.1:p.Val168SerfsTer28 | p.Val168SerfsTer28 | LP | frameshift variant | 3 |

*All non-autosomal dominant or X-linked pathogenic mutations in the PID gene set previously reported in the Clinvar database. AR, autosomal recessive. P, pathogenic. LP, likely pathogenic.*

**Supplementary table S10. Clinvar GWAS Candidate non-AD mutations**

| Gene | Inherit-ance | Mutation | Clinvar | CAPICE score | genomic mutation location | protein change | VIP annot. | Mutation Type | Times detected |
|---|---|---|---|---|---|---|---|---|---|
| *IL7R* | AR | c.470del | P | 0,54579556 | NP 002176.2:p.Lys157ArgfsTer4 | p.Lys157ArgfsTer4 | LP | frameshift variant | 10 |
| *SLC22A5* | AR | c.1219G>T | LP | 0,9974833 | NP 001295051.1:p.Glu407Ter | p.Glu407Ter | LP | stop gained | 1 |

*All non-autosomal dominant or X-linked pathogenic mutations in the IBD GWAS Candidate gene set previously reported in the Clinvar database. AR, autosomal recessive. P, pathogenic. LP, likely pathogenic.*

**Supplementary table S11**

**All VKGL mutations (see Excel file)**

**Supplementary table S12**

**All Clinvar mutations (see Excel file)**

**Supplementary table S13**

**VEO-IBD VKGL (see Excel file)**

**Supplementary table S14**

**VEO-IBD Clinvar (see Excel file)**

**Supplementary table S15**

**GWAS Candidate VKGL (see Excel file)**

**Supplementary table S16**

**GWAS Candidate Clinvar (see Excel file)**

**Supplementary table S17**

**GWAS Loci VKGL (see Excel file)**

**Supplementary table S18**

**GWAS Loci Clinvar (see Excel file)**

**Supplementary table S19**

**PID VKGL (see Excel file)**

**Supplementary table S20**

**PID Clinvar (see Excel file)**

**Supplementary table S21**

**GWAS Candidate all LP (see Excel file)**

**Supplementary table S22**

**GWAS Candidate AD (see Excel file)**

**Supplementary table S23**
**GWAS Candidate AR (see Excel file)**

**Supplementary table S24**
**GWAS Loci All LP (see Excel file)**

**Supplementary table S25**
**GWAS Loci AD (see Excel file)**

**Supplementary table S26**
**GWAS Loci AR (see Excel file)**

**Supplementary table S27**
**GWAS Loci X-linked males (see Excel file)**

**Supplementary table S28**
**PID All LP (see Excel file)**

**Supplementary table S29**
**PID AD (see Excel file)**

**Supplementary table S30**
**PID AR (see Excel file)**

**Supplementary table S31**
**PID X-linked males (see Excel file)**

**Supplementary table S32**
**VEO-IBD All LP (see Excel file)**

**Supplementary table S33**
**VEO-IBD AD (see Excel file)**

**Supplementary table S34**
**VEO-IBD AR (see Excel file)**

**Supplementary table S35**
**VEO-IBD X-linked males (see Excel file)**

**Supplementary table S36**
**Mutations in previously identified genes (see Excel file)**

**Supplementary figure S1**

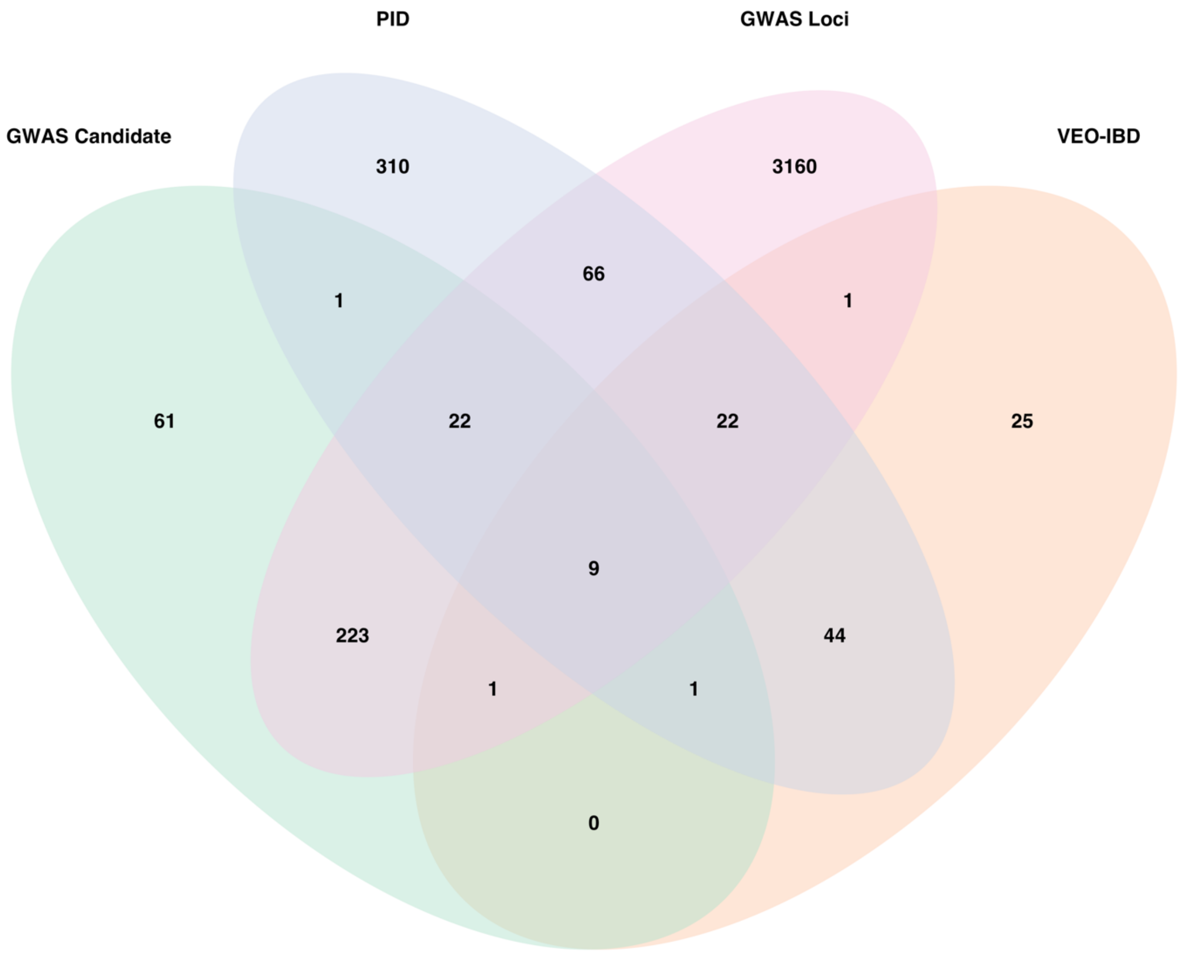


**Supplementary figure S1. Overlap between gene sets.** Overlap between the four gene sets is shown. GWAS Candidate genes are as defined by Huang et al (2021). Primary Immune Deficiency genes (PID) and very early onset IBD genes (VEO-IBD) are as defined by the UMCG Diagnostics lab. GWAS Loci genes contain all genes in the loci defined by the GWAS studies.

**Supplementary figure S2**

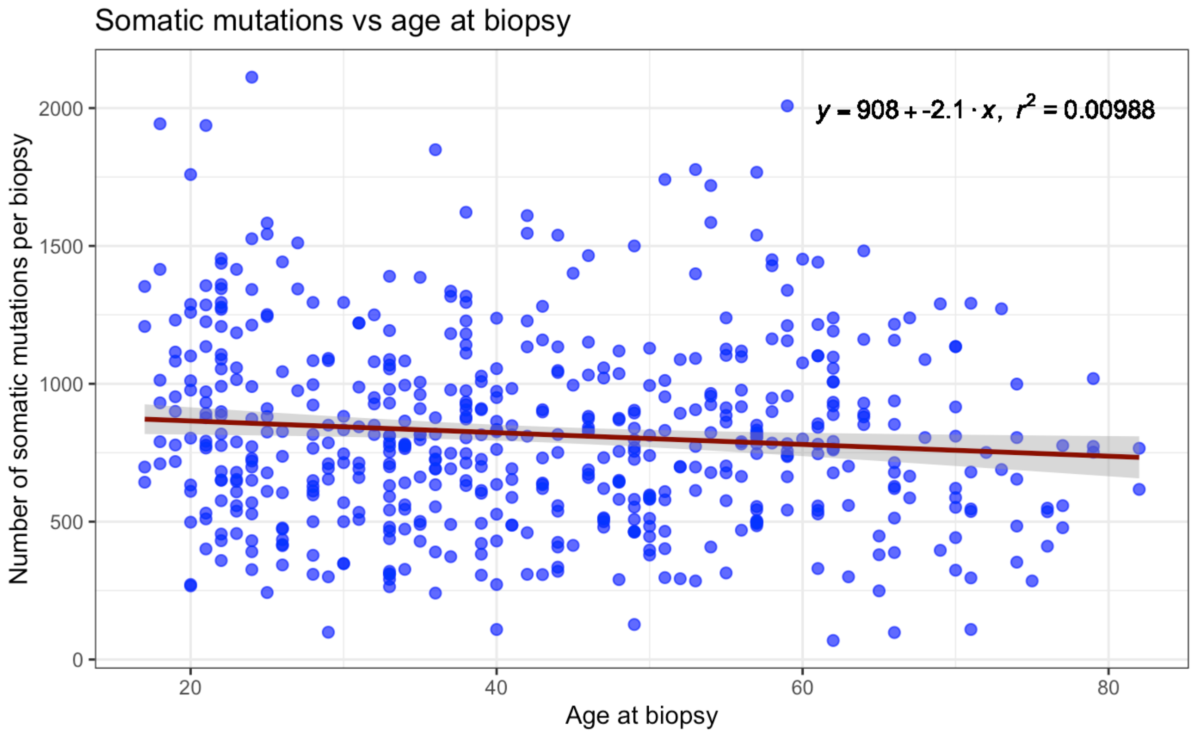


***Supplementary figure S2. Correlation of somatic mutations versus age at biopsy***. *Shown are the number of mutations per biopsy versus age at moment of biopsy. A small negative correlation is observed.*

**Supplementary figure S3**

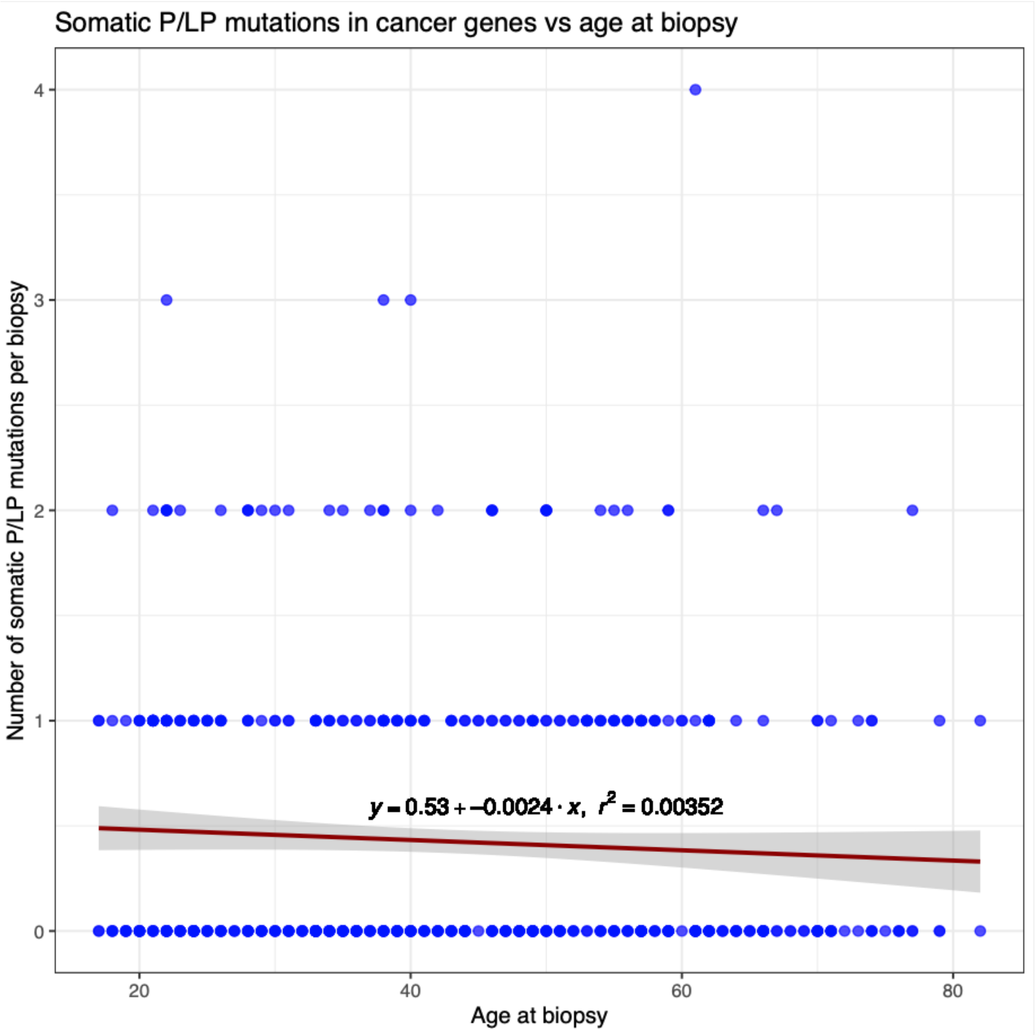


***Supplementary figure S3. Correlation of Pathogenic/Likely Pathogenic (P/LP) somatic mutations in cancer genes versus age at biopsy****. Shown are the number of P/LP mutations in cancer genes per biopsy versus age at moment of biopsy. A small negative correlation is observed.*

**Supplemental figure S4**

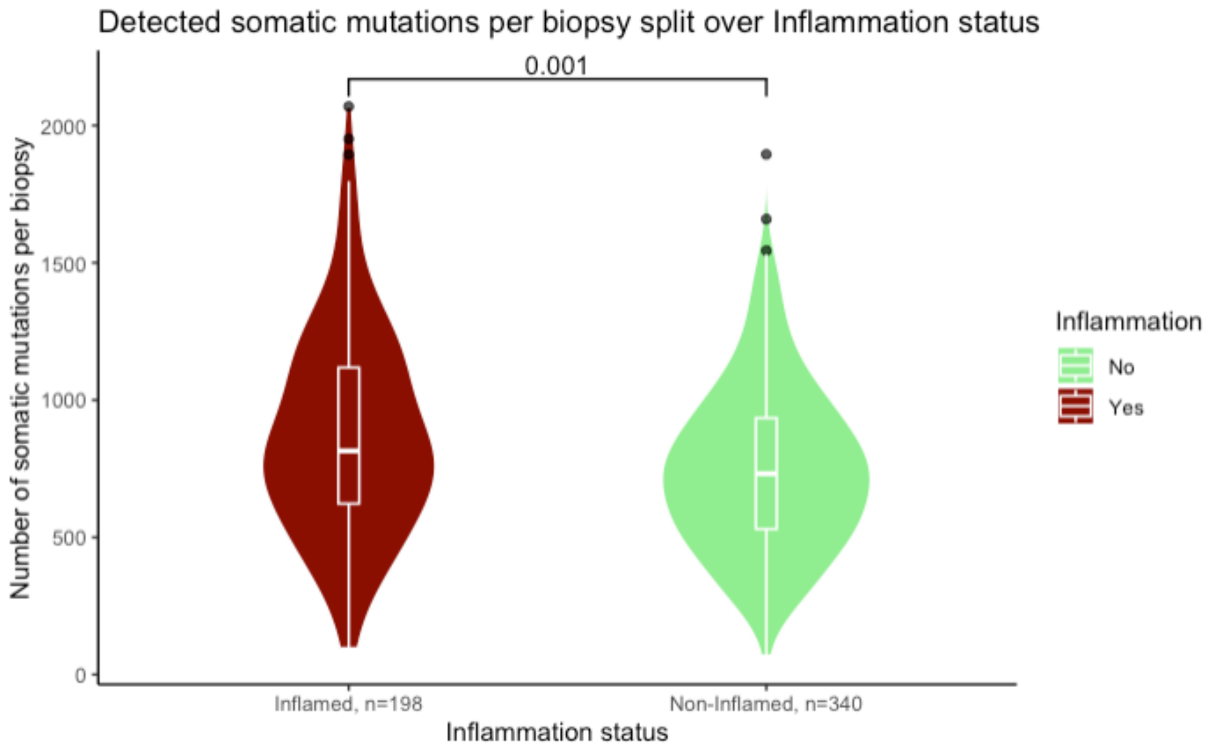


***Supplementary figure S4. Somatic mutations versus inflammation status of the tissue of origin per biopsy.*** *Shown are the number of somatic mutations observed per biopsy, grouped by inflammation status of the tissue. A significant increase of somatic mutations is observed under inflamed conditions (Mann-Whitney U, p=0.001).*

**Supplemental figure S5**

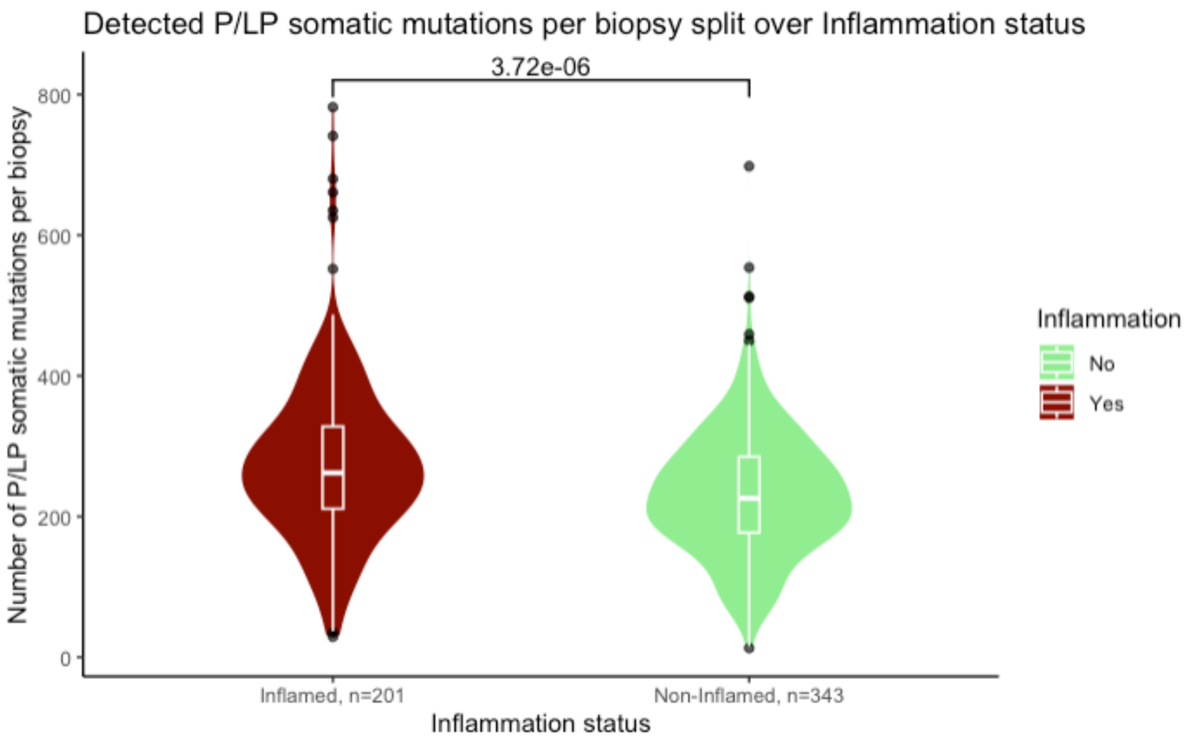


***Supplementary figure S5. Likely pathogenic/Pathogenic (LP/P) somatic mutations versus inflammation status of the tissue of origin per biopsy.*** *Shown are the number of LP/P somatic mutations observed per biopsy, grouped by inflammation status of the tissue. A significant increase of LP/P somatic mutations is observed under inflamed conditions (Mann-Whitney U, p=3.74e-06).*

**Supplemental figure S6**

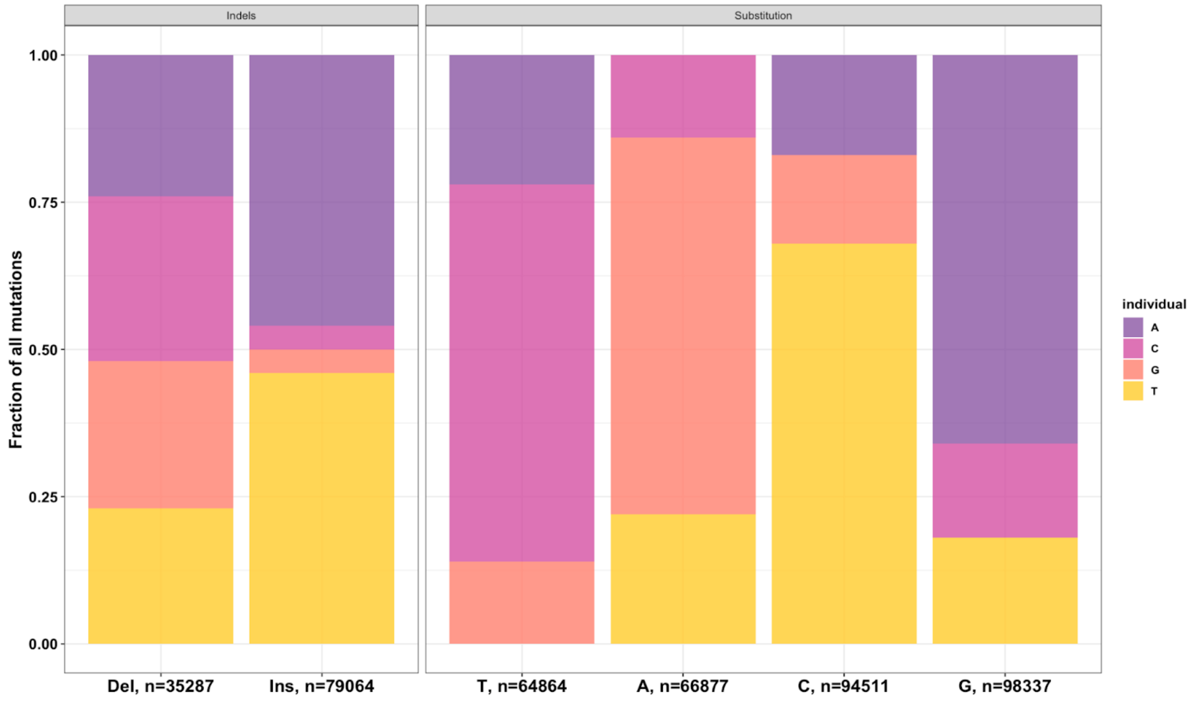


***Supplementary figure S6. Somatic mutations are not equally distributed over DNA base types.*** *We investigated the likelihood of specific base changes in the somatic mutations data. We detected 94,511 C-substitutions (C>T most common) and 98,337 G-substitutions (G>A most common). We observed 66,877 A-substitutions (A>G most common) and 64,864 T-substitutions (T>C most common). We also observed 79,064 insertions and 35,287 deletions. Insertions were primarily A and T. Deletions were observed in similar ratios across all bases.*
